# Sex-stratified and interaction multi-omics analysis of DNA methylation and gene expression in asthma

**DOI:** 10.64898/2026.10.19.26365928

**Authors:** Eskezeia Y. Dessie, Yadu Gautam, Bradley H. Rosen, Angelico Mendy, Mingon Kang, Tesfaye B. Mersha

## Abstract

**Background:** Sex differences in asthma are well recognized, but sex-associated airway DNA methylation (DNAm) patterns and their relationships with gene expression remain incompletely characterized. We therefore conducted cis- and trans-expression quantitative trait methylation (eQTM) analyses using sex-stratified and CpG-by-sex interaction models.

**Methods:** We analyzed nasal airway epithelial DNAm and transcriptomic data from 298 participants in the CAAPA discovery cohort. Asthma-associated differentially methylated CpGs (DMCs) were identified using sex-combined and sex-stratified models. Cis- and trans-eQTM analyses were then performed in participants with asthma using sex-combined, sex-stratified, and CpG-by-sex interaction models. To prioritize asthma-associated regulatory relationships, eQTMs that were also significant in non-asthmatic controls were excluded from downstream analyses. Replication in an independent bronchial epithelial cohort was defined by concordant effect direction and P < 0.05. Replicated targets underwent pathway and network analyses.

**Findings:** Asthma-associated DMCs were predominantly hypomethylated in females and hypermethylated in males. Among trans-eQTMs, 379 associations were shared across all four models, whereas the majority were model-specific, indicating extensive sex-dependent regulatory architecture. We replicated 3, 1, 9, and 2 cis-eQTMs and 96, 84, 216, and 41 trans-eQTMs in the sex-combined, female-stratified, male-stratified, and interaction models, respectively. Sex-combined targets implicated innate immune and inflammatory signaling (*TNF*, *IRF8*); female-stratified targets implicated cytoskeletal remodeling, focal adhesion, and PI3K-Akt signaling (*ITGA10*); male-stratified targets implicated transcriptional regulation and post-translational modification (*MOV10*, *HDAC7*, *CSF1*, *KAT2A*); and interaction targets implicated lipid-mediator metabolism and inflammatory signaling and resolution (*CYP2C9*, *LTB4R*, *CCL19*). Our sex-dependent analyses showed that the majority of eQTMs resided within the trans-regulatory architecture, suggesting that sex-dependent gene expression differences are associated predominantly with trans rather than cis regulatory mechanisms. Pathway analyses revealed enrichment of innate immunity in the sex-combined model, structural remodeling in females, transcriptional control in males, and inflammatory resolution in the sex-interaction model.

**Interpretation:** Sex-specific and sex-dependent airway methylation-expression associations were largely undetected by sex-combined analysis. Sex-aware analytical models may help resolve epigenetic mechanisms underlying sex differences in asthma biology.

**Research in context Evidence before this study:** Asthma shows well-recognized sex differences in prevalence, severity, and clinical outcomes, yet the molecular mechanisms underlying these differences remain incompletely understood. Airway DNAm has been implicated in asthma pathogenesis and may contribute to sex-related disease heterogeneity. However, most epigenomic studies have analyzed females and males together, adjusting for sex as a covariate, an approach that may leave sex-specific and sex-dependent methylation-expression relationships undetected. Consequently, whether airway DNA methylation regulates gene expression differently in females and males, through cis- or trans-acting mechanisms, remains largely unknown.

**Added value of this study:** We integrated airway epithelial DNAm and transcriptomic data to perform cis- and trans-eQTM analyses using sex-combined, sex-stratified, and CpG-by-sex interaction models. Asthma-associated CpGs showed distinct sex-specific methylation patterns, with predominantly hypomethylated CpGs in females and hypermethylated CpGs in males. Most eQTMs were model-specific, with only 379 trans-eQTMs shared across all analytical models, indicating extensive sex-dependent regulatory architecture. Replication in an independent bronchial epithelial cohort confirmed both shared and sex-specific eQTMs, with a substantially larger number of trans than cis associations. Replicated targets identified model-specific biological pathways, including innate immunity and inflammatory signaling in the sex-combined model, structural remodeling in females, transcriptional regulation in males, and lipid-mediator metabolism and inflammatory resolution in the sex-interaction model. Importantly, many of these regulatory relationships were not identified by sex-adjusted analyses.

**Implications of all the available evidence:** Our findings demonstrate that sex-specific and sex-dependent methylation-expression associations represent an important component of airway epigenetic regulation in asthma and are largely undetected by sex-adjusted analyses. Incorporating sex-aware analytical frameworks into epigenomic and transcriptomic studies may improve the identification of biologically relevant regulatory mechanisms, sex-specific biomarkers, and therapeutic targets. Replication of key findings in bronchial epithelium further supports their biological relevance and potential translational value for precision medicine approaches in asthma.

**Graphical Abstract:** 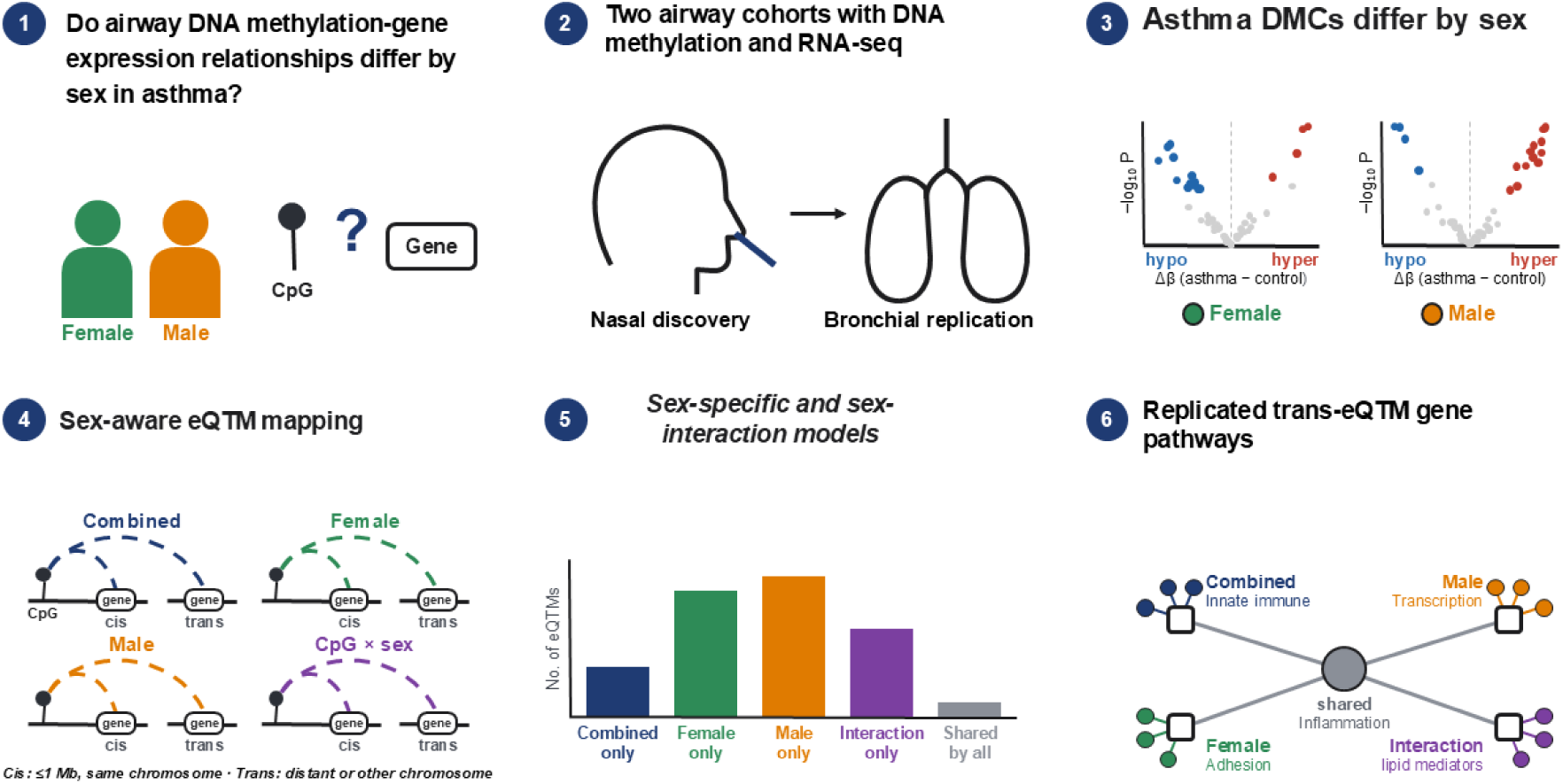

## Background

Asthma affects more than 260 million people worldwide^1^, and displays marked sexual dimorphism. Disease prevalence and severity shift from male predominance in childhood to female predominance after puberty^2–4^. These sex-specific differences are likely driven by complex interactions among genetic, epigenetic, immunological, hormonal, and environmental factors that shape airway inflammation and immune function^5–7^. Despite these well-established epidemiological patterns, how biological sex shapes asthma susceptibility and heterogeneity at the molecular level remains poorly understood, and the contribution of sex-specific epigenetic regulation to asthma pathogenesis has not been systematically examined^3,7^.

The airway epithelium is a primary site of asthma pathobiology. DNA methylation is a cell-type-specific epigenetic mark associated with gene expression^8^, and it reflects both genetic and environmental influences^9^. Epigenome-wide association studies (EWAS) have identified reproducible asthma-associated methylation in nasal airway epithelium^10^. Sex hormones also shape DNA methylation: estrogen and progesterone influence DNA methyltransferase activity^11^, and testosterone is inversely associated with global methylation^12^. Sex-associated methylation differences at asthma-related inflammatory genes suggest a potential role for airway epigenetic regulation in asthma sexual dimorphism^3,13,14^. Such effects would be expected to manifest through sex-specific relationships between DNA methylation and gene expression.

Despite growing evidence for sex-specific epigenetic regulation in asthma, several key knowledge gaps remain. First, CpG sites identified through epigenome-wide association studies (EWAS) are typically assigned to the nearest gene, although this assignment may not reflect their true regulatory targets. Expression quantitative trait methylation (eQTM) analysis directly links DNA methylation to gene expression, yet asthma studies have focused largely on cis associations^15^. This focus may overlook substantial regulatory activity, as many expression-associated CpGs in airway epithelium regulate genes at distal loci^16^, potentially through transcription factor-mediated networks^17^. Second, airway eQTM studies, like large multi-tissue resources^18^, have modeled sex only as a covariate. Sex-combined models can obscure effects that differ by sex; methylation at cg20891917 (IFRD1), for example, shows opposite associations with asthma in females and males^3^. Stratified analyses alone also cannot distinguish true sex differences from differences in statistical power, which requires formal interaction testing. Third, nasal findings are rarely tested in lower-airway tissue, although nasal and bronchial epithelia share much of their asthma-related expression^19,20^.

Here, we tested the hypothesis that methylation–expression relationships in the asthmatic airway epithelium differ by sex, and that sex-aware models detect associations missed by sex-combined analysis. Using nasal airway epithelium, we identified sex-combined and sex-stratified asthma-associated differentially methylated CpGs. We then mapped cis- and trans-eQTMs in participants with asthma using sex-combined, female-stratified, male-stratified and CpG×sex interaction models, and related eQTM target genes to differential expression. Finally, we tested associations in an independent bronchial epithelial cohort and examined the biological pathways implicated by replicated targets.

## Methods

### Discovery dataset

The discovery dataset comprised 298 participants (183 females and 115 males) from the Consortium on Asthma among African Ancestry Populations (CAAPA), a multicenter cohort including participants from U.S. and international sites. Paired nasal airway epithelial DNA methylation (GEO: GSE250513) and RNA sequencing (RNA-seq; GEO: GSE240567) data were available for all participants. This included 136 asthma cases (81 females, 55 males) and 162 controls (102 females, 60 males). Methylation was profiled on the Illumina EPIC array and processed using minfi^21^; matched RNA-seq (GSE240567) underwent standard alignment^22^, quantification^23^, low-count filtering, and TMM-based normalization^24^. Detailed recruitment criteria, QC thresholds, and retained feature counts are provided in the original study^25^ and Supplementary Methods.

### Differential methylation analysis

To prioritize asthma-associated DNAm sites for downstream regulatory analyses, we performed case-control differential methylation analyses using three parallel models: sex-combined, female-specific, and male-specific. DNA methylation (DNAm) sites with low (mean β < 0.2) or high (mean β > 0.8) methylation were retained and further restricted to sites showing an absolute difference in mean methylation between participants with asthma and controls of |Δβ| > 0.01. Differential methylation was assessed using limma linear models adjusted for age, ancestry, site, and array plate, with additional adjustment for sex in the combined analysis. DNAm sites meeting |Δβ| > 0.01 and nominal P < 0.05 were retained as differentially methylated CpGs (DMCs) for downstream regulatory analyses and classified as hypermethylated or hypomethylated according to the direction of Δβ. The nominal significance threshold was used as a screening criterion to prioritize potentially asthma-associated DNAm sites for subsequent regulatory analyses.

### Cis- and trans-eQTM models

We used regression models to investigate associations between DNA methylation and gene expression in subjects with asthma. CpG–gene associations were examined using four eQTM models: sex-combined, female-specific, male-specific, and CpG × sex interaction models, with CpGs selected from the corresponding DMCs identified in the upstream differential methylation analyses. Within each model, cis- and trans-eQTM analyses were performed to characterize local and distal methylation-gene expression associations. Collectively, these models were designed to identify shared, sex-specific, and sex-dependent methylation-gene expression relationships and to characterize their potential regulatory relevance in asthma. Detailed descriptions of each eQTM model are provided below.

1. Sex-combined model Expression*_i_* = *β*_0_ + *β*_1_ CpG*_i_* + ω*^T^ Cov_i_* + *∊_i_*, where includes sex, age, site, plate, PC1, and PC2. CpGs were selected from the sex-combined differential methylation analysis using a nominal *P* < 0.05 and subsequently evaluated for cis- and trans-eQTM associations. The CpG coefficient (*β*_1_) estimates the adjusted methylation–gene expression association across all samples with asthma.
2. Female-specific model Expression*_i_* = *β*_0_ + *β*_1_ CpG*_i_* + *ω^T^ Cov_i_* + *∊_i_* , where *β*_0_ is the intercept, *β*_1_ represents the association between CpG methylation and gene expression, *Cov_i_* denotes the vector of covariates (age, site, plate, PC1, and PC2), *ω* represents their corresponding regression coefficients, and *∊_i_* is the residual error. The analysis was performed in female subjects, and CpGs were selected from the female-specific differential methylation analysis using a nominal *P* < 0.05. Therefore *β*_1_ estimates the effect size of the adjusted methylation–gene expression association among females. This model was used to identify cis- and trans-eQTM associations detectable specifically in the female-stratified analysis, including associations not detected in the male-specific or sex-combined analyses.
3. Male-specific model Expression*_i_* = *β*_0_ + *β*_1_ CpG*_i_* + *ω^T^ Cov_i_* + *∊_i_*, where is the intercept, represents the association between CpG methylation and gene expression, *Cov_i_* denotes the vector of covariates (age, site, plate, PC1, and PC2), *ω* represents their corresponding regression coefficients, and*∊_i_* is the residual error. The analysis was performed in male subjects, and CpGs were selected from the male-specific differential methylation analysis using a nominal *P* < 0.05. Therefore, *β*_1_ estimates the effect size of the adjusted methylation–gene expression association among males. This model was used to identify cis- and trans-eQTM associations detectable specifically in the male-stratified analysis, including associations not detected in the female-specific or sex-combined analyses.
4. Sex interaction model Expression*_i_* = *β*_0_ + *β*_1_CpG*_i_* + *β*_2_Sex*_i_* + *β*_3_ (CpG*_i_* × Sex*_i_*) *ω^T^ Cov_i_* + *∊_i_*, where *β*_0_ is the intercept; *β*_1_ represents the CpG methylation–gene expression association; *β*_2_ represents the main effect of sex; and *β*_3_ represents the CpG × sex interaction effect, testing whether the methylation–gene expression association differs between males and females. *Cov_i_* includes age, site, plate, PC1, and PC2, with corresponding coefficients represented by *ω*. The significance of the CpG × sex interaction was examined using a likelihood-ratio test comparing models with and without the interaction term. Significant interaction eQTMs therefore identified CpG–gene associations whose magnitude and/or direction differed by sex.

All four models were run separately for cis (CpG–gene pairs within 1 Mb of the TSS) and trans (>1 Mb or different chromosome) associations using MatrixEQTL^26^. Methylation and gene expression values were standardized to a mean of 0 and standard deviation of 1 before model fitting to comparison of effect estimates across models. Covariates included age, study site, array plate, and the first two ancestry principal components, with sex additionally included as a covariate in the sex-combined model. For cis-eQTM analyses, statistical significance in the primary discovery analysis was defined using a Benjamini-Hochberg false discovery rate (FDR) < 0.05. For trans-eQTM analyses, significant associations were defined using FDR < 0.05 to account for the substantially greater multiple-testing burden associated with genome-wide trans CpG–gene testing, consistent with thresholds used in previous large-scale airway eQTM studies^16^.

### Differential expression analysis and integration with eQTM target genes

To evaluate the functional relevance of methylation-regulated genes, we performed differential gene expression analyses using normalized RNA-seq data from the CAAPA cohort (GEO: GSE240567). Gene expression was compared between participants with asthma and non-asthmatic controls separately in females and males. Linear models with empirical Bayes moderation were fitted using the limma package. Sex-stratified models were adjusted for age, ancestry, site, and array plate. For the targeted evaluation of replicated eQTM target genes, a nominal threshold of P < 0.05 was used to identify differentially expressed genes (DEGs), as these genes were supported by independent cis- and trans-eQTM methylation-expression associations. Cis- and trans-eQTM target genes and DEGs were reported separately, and their overlap was subsequently evaluated to identify methylation-associated transcriptional alterations in asthma within each sex. Replicated cis- and trans-eQTM target genes were carried forward for downstream pathway enrichment and network analyses. Genes exhibiting both methylation-expression associations and differential expressions were highlighted as candidate methylation-regulated asthma genes. All analyses were performed in R (version 4.5.1).

### Replication dataset

We sought to replicate our discovery eQTM findings using an independent bronchial airway epithelial dataset with paired DNA methylation and gene expression data generated from bronchial epithelial samples collected via bronchoscopy (Illumina HumanMethylation450 methylation data, GSE85568; RNA-seq data, GSE85567)^27^. The replication dataset comprised 81 adults (56 females and 25 males), including 54 individuals with asthma (39 females and 15 males) and 27 controls (17 females and 10 males). For cross-platform replication, analyses were restricted to CpG sites represented on both the Illumina EPIC and HumanMethylation450K arrays. Within each dataset, sex-combined, female-specific, and male-specific differential methylation analyses were performed, and CpGs showing nominal evidence of methylation difference between participants with asthma and controls (|Δβ| > 0) and nominal P < 0.05 were used for eQTM analysis. We applied the same four-model eQTM framework used in the discovery analysis, including sex-combined, female-specific, male-specific, and sex-interaction models, separately to cis and trans CpG–gene pairs. All models were adjusted for age, and the sex-combined and sex-interaction models were additionally adjusted for sex. The replication analyses were performed for the CpG-gene pairs that were significant in the discovery dataset. Replication significance was defined at nominal significance (P < 0.05) in the bronchial airway epithelial replication cohort and showed an effect direction concordant with that observed in the nasal airway epithelial discovery cohort^15^.

### Pathway Enrichment of eQTM Derived Gene Signatures

We used GSEA (v4.4.0)^28^ independently to gene lists from replicated four sex-stratified eQTM models (sex-combined, female-specific, male-specific, sex-interaction), ranking genes by −log₁₀ p-value. We evaluated enrichment against curated canonical pathway collections (MSigDB 2026.1)^29^ using 1,000 permutations, gene set size 15–500, and FDR < 0.05. We integrated leading-edge genes and their pathway memberships across models into an undirected bipartite graph using the igraph R package, with edges denoting leading-edge co-membership in ≥1 significant pathway. We define hub genes as those connected to ≥2 distinct significant pathways (node degree ≥2) and multi-model genes as those identified in ≥2 models. We visualized the network using force-directed layout, with node size scaled proportionally to degree.

## Results

### Characteristics of the study population

The nasal airway epithelial discovery cohort included 298 participants, comprising 115 males (48% with asthma) and 183 females (44% with asthma). Age did not differ significantly between participants with asthma and controls among either males (23.5 ± 16.6 vs. 25.2 ± 14.6 years, *P* = 0.551) or females (31.0 ± 18.2 vs. 28.1 ± 15.7 years, *P* = 0.264). The bronchial airway epithelial replication cohort included 81 participants, comprising 25 males (60% with asthma) and 56 females (70% with asthma). Age was comparable between male participants with asthma and controls (36.7 ± 14.3 vs. 39.6 ± 12.3 years, *P* = 0.589), whereas female participants with asthma were older than female controls (41.5 ± 12.3 vs. 34.6 ± 10.9 years, *P* = 0.046) (Table 1).

**Table 1.**
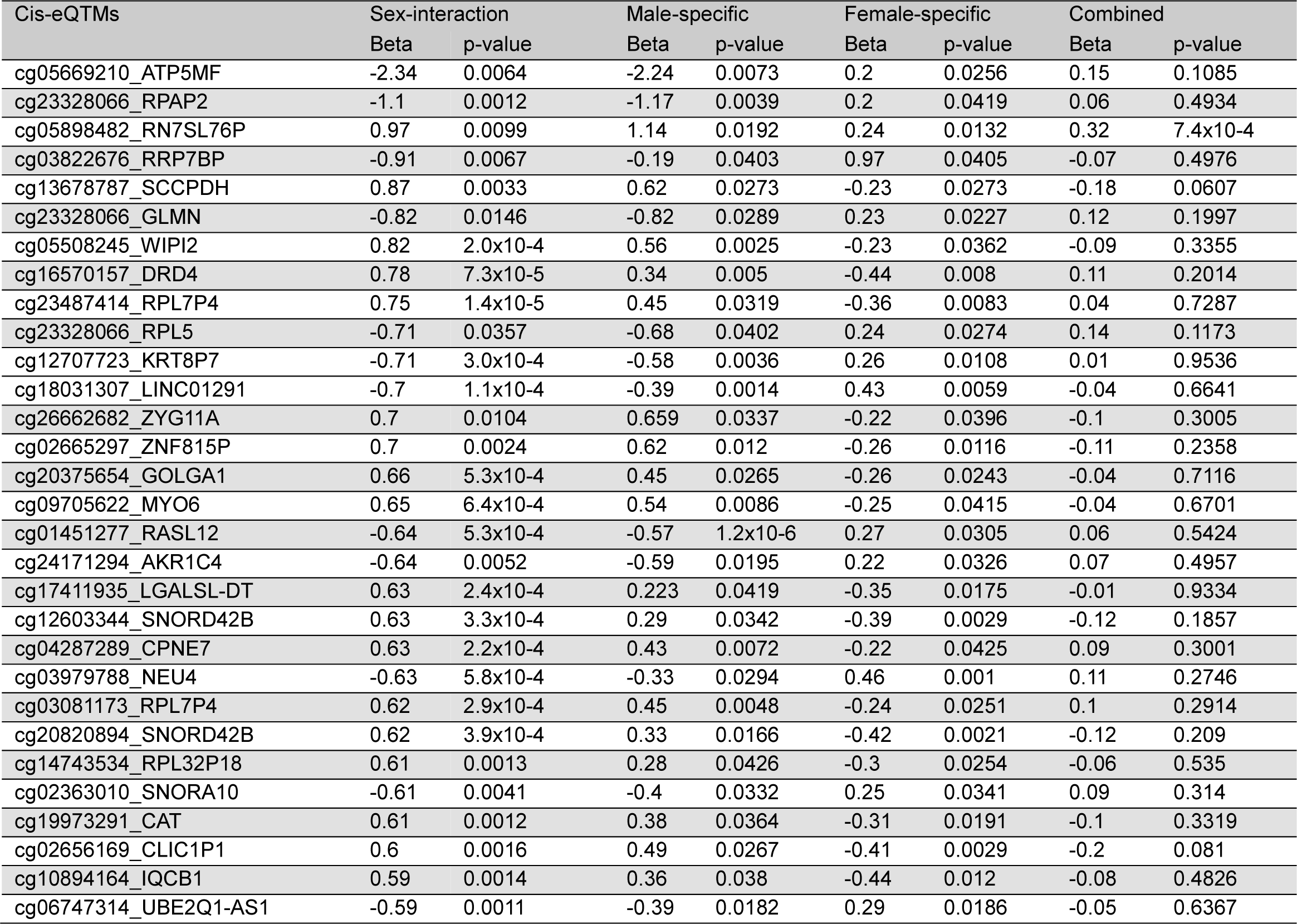
Dataset characteristics by sex and disease status. Baseline characteristics of discovery (n = 298) and replication (n = 81) datasets. Data means SD or n (%). P-values: t-test (age) or χ² test comparing controls vs. asthma within each sex. Bold indicates P < 0.05.

### DNAm signatures reveal shared and sex-specific epigenetic associations

To delineate the epigenetic landscape associated with asthma and its dependence on sex, we performed differential methylation analyses comparing participants with asthma and controls in the sex-combined cohort (n = 298; 136 asthma and 162 controls) and separately in females (n = 183) and males (n = 115). Across 834,663 CpGs, DNAm sites with low (mean β < 0.2) or high (mean β > 0.8) methylation, an absolute mean methylation difference of |Δβ| > 0.01 between participants with asthma and controls, and a limma-derived unadjusted P < 0.05 were defined as differentially methylated CpGs (DMCs). We identified 5,331 differentially methylated CpGs (DMCs) in the sex-combined analysis, comprising 2,587 hypermethylated and 2,744 hypomethylated CpGs (Fig. 1a). Sex-stratified analyses identified 3,054 DMCs in females and 2,729 DMCs in males. Female-associated DMCs were predominantly hypomethylated (n = 2,205; 72.2%), with 849 (27.8%) hypermethylated CpGs, whereas male-associated DMCs showed the opposite pattern, with 1,693 (62.0%) hypermethylated and 1,036 (38.0%) hypomethylated CpGs. These results reveal distinct sex-dependent DNAm signatures of asthma, characterized predominantly by hypomethylation in females and hypermethylation in males.

**Figure 1.**
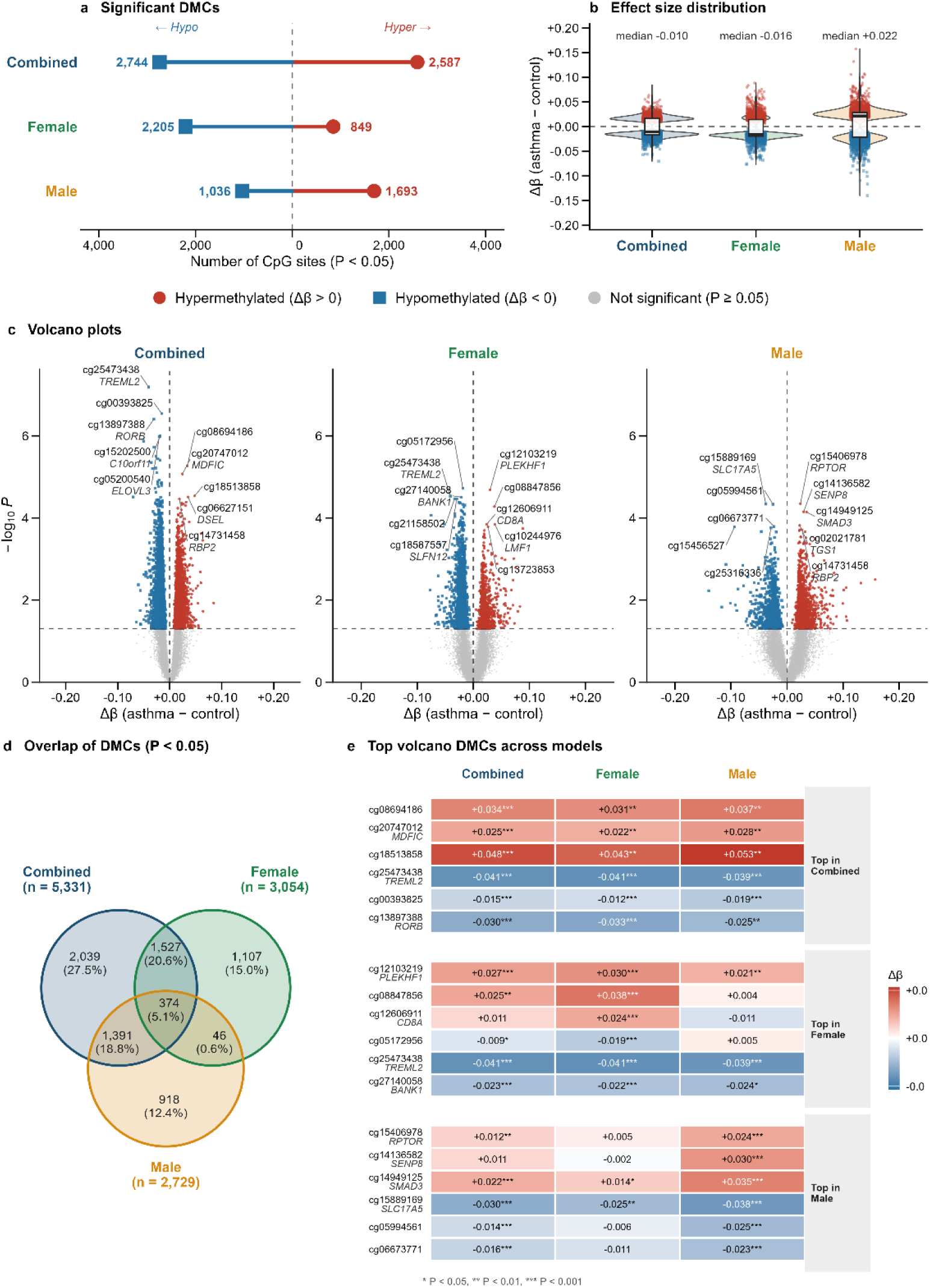
Differentially methylated CpGs (DMCs) associated with asthma in combined and sex-stratified analyses. DNA methylation was compared between participants with asthma and controls using covariate-adjusted limma models. DMCs were defined as CpGs with nominal *P* < 0.05. Red and blue indicate hypermethylated and hypomethylated CpGs, respectively; navy, green, and orange represent combined, female, and male analyses. (a) Numbers of hypo- and hypermethylated DMCs identified in the combined, female, and male analyses. (b) Distribution of methylation differences (Δβ = mean β-asthma − mean β-control) among DMCs. Violin plots show density distributions, with medians and interquartile ranges indicated. (c) Volcano plots showing Δβ versus −log10(*P*) for all tested CpGs. The horizontal dashed line denotes *P* = 0.05 and the vertical dashed line denotes Δβ = 0. The five most significant hypermethylated and hypomethylated CpGs are labeled. (d) Venn diagram showing overlap of DMCs across combined, female, and male analyses. (e) Cross-analysis comparison of the top hypermethylated and hypomethylated CpGs from each analysis. Heatmap colors represent Δβ values, and asterisks indicate significance (*P* < 0.05, \**P* < 0.01, \*\**P* < 0.001).

The distribution of effect sizes also differed between sexes (Fig. 1b). DMCs in the female analysis had a median Δβ of −1.63% (range, −7.69% to +8.84%), whereas DMCs in the male analysis had a median Δβ of +2.20% and a wider range (−13.98% to +15.74%). Male effect sizes were larger compared to female. Volcano plots of the three analyses (Fig. 1c) show the most significant associations. In the sex-combined analysis, the top CpGs were cg25473438 (*TREML2*; P = 6.4 × 10⁻׈ and cg13897388 (*RORB*; P = 3.9 × 10In females, the top CpGs were cg12103219 (*PLEKHF1*; P = 2.1 × 10⁻׈) and cg25473438 (*TREML2*; P = 2.9 × 10⁻׈). In males, the top CpGs were cg15406978 (*RPTOR*; P = 4.5 × 10⁻׈) and cg15889169 (*SLC17A5*; P = 4.6 × 10⁻׈).

DMCs overlapped only partially across analyses (Fig. 1d; Supplementary Table S1a–c). Of 7,402 unique DMCs, 374 were shared by all three analyses, and a further 46 were shared by the female and male analyses only. Most sex-stratified DMCs were therefore found in one sex only: 1,107 in females only and 918 in males only. Effect sizes comparison of across analyses for the leading CpGs of each model are shown (Fig. 1e). The top sex-combined CpGs showed concordant methylation direction and similar magnitude in both sexes. For example, cg25473438 (*TREML2*) had similar direction in females (P = 2.9 × 10⁻׈) and in males (P = 9.0 × 10⁻׈). In contrast, several top sex-stratified CpGs showed effects largely confined to one sex.

These included cg12606911 (*CD8A*) in females (P = 1.4 × 10⁻׈; males P = 0.38), and cg14136582 (*SENP8*) and cg15406978 (*RPTOR*) in males (females P = 0.35 and 0.57).

Together, these findings indicate both shared asthma-associated methylation changes and sex-specific DMCs that were not identified in the sex-combined analysis. We therefore used DMCs identified in each analysis for subsequent cis- and trans-eQTM analyses to determine whether these asthma-associated methylation differences were associated with gene expression.

### Cis- and trans-eQTM analysis

To examine asthma-associated CpGs influence gene expression, we conducted cis- and trans-eQTM analyses in asthma cases from the nasal cohort using four models: sex-adjusted combined, female-specific, male-specific, and sex-interaction. Associations also identified in control-only analyses were removed to identify on asthma-specific regulatory associations. The numbers of cis- and trans-eQTMs identified in each model are presented in Fig. 2a, b. Sex-stratified analyses uncovered more trans-eQTMs associations than the sex-adjusted combined model, suggesting that sex-specific regulatory effects contribute substantially to the epigenetic architecture of asthma. Examination of cis- and trans-eQTM effect-size distributions showed greater dispersion in the male-specific and sex-interaction models than in the sex-adjusted combined model (Fig. 2 c, d). Detailed findings from each model are presented below.

**Figure 2.**
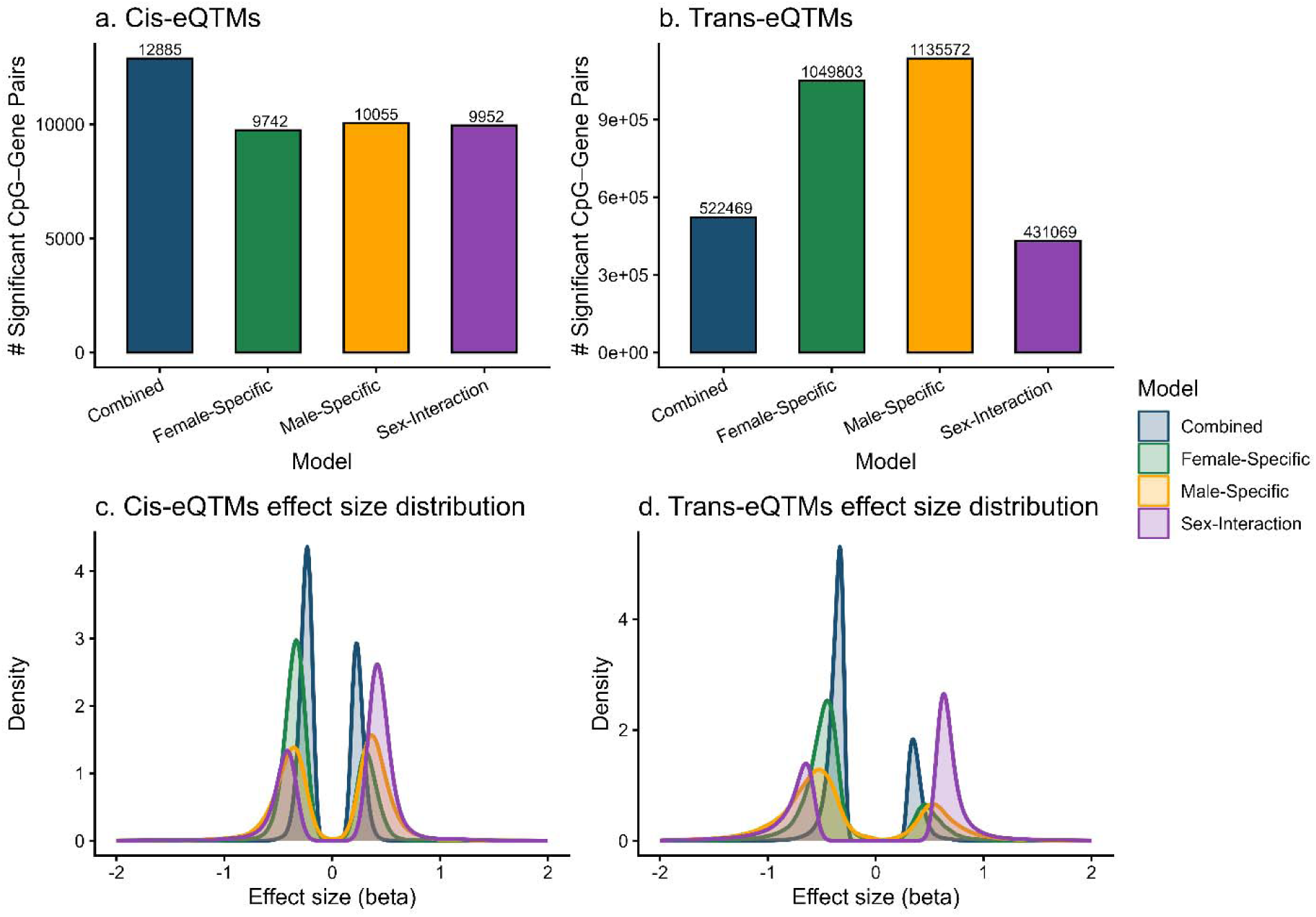
Asthma-specific eQTM associations across sex-stratified models. a) Number of asthma-specific cis-eQTM pairs (CpG–gene pairs within 1 Mb) identified in each sex-stratified model. Asthma-specific pairs were defined as associations reaching nominal significance in asthma cases (p < 0.05) but not in control subjects (p ≥ 0.05). b) Number of asthma-specific trans-eQTM pairs (CpG–gene pairs beyond 1 Mb) per model, defined as associations reaching FDR < 0.05 in asthma cases but not in control subjects (FDR ≥ 0.05). c) Density distribution of effect sizes (β) for asthma-specific cis-eQTM association across models. d) Density distribution of effect sizes (β) for asthma-specific trans-eQTM association across models. In all panels, colors indicate the sex-stratified model: navy = Combined, green = Female-specific, orange = Male-specific, purple = Sex-interaction. Effect sizes represent standardized regression coefficients from linear models of CpG methylation on gene expression. Abbreviations: β, standardized regression coefficient; cis-eQTM, expression quantitative trait methylation locus within 1 Mb; trans-eQTM, expression quantitative trait methylation locus beyond 1 Mb; FDR, false discovery rate.

#### Sex-combined model

In the sex-adjusted combined analysis, 12,885 cis-eQTMs were identified at P < 0.05, of which 88 remained significant at FDR < 0.05 (Fig. 2a; Supplementary Table S2a). In addition, 522,469 trans-eQTMs were significant at FDR < 0.05 (Fig. 2b; Supplementary Table S2b).

#### Female model

In the female-stratified analysis, 9,742 cis-eQTMs were identified at P < 0.05, of which 88 remained significant at FDR < 0.05 (Fig. 2a; Supplementary Table S2c). In addition, 1,053,891 trans-eQTMs were significant at FDR < 0.05, 823,286 of which were identified only in the female model ((Fig. 2b; Supplementary Table S2d). Ranked by P value, the top cis-eQTM genes were *OR2T12, RN7SKP192*, *LINC02829*, *BMP6P1, IGF2-AS and KRT17P7*, and the top trans-eQTM genes were *RN7SL583P, ASB17, HMGB1P18, PRELID1P7* and *CRIP1P2* (Fig. 3a-d).

**Figure 3.**
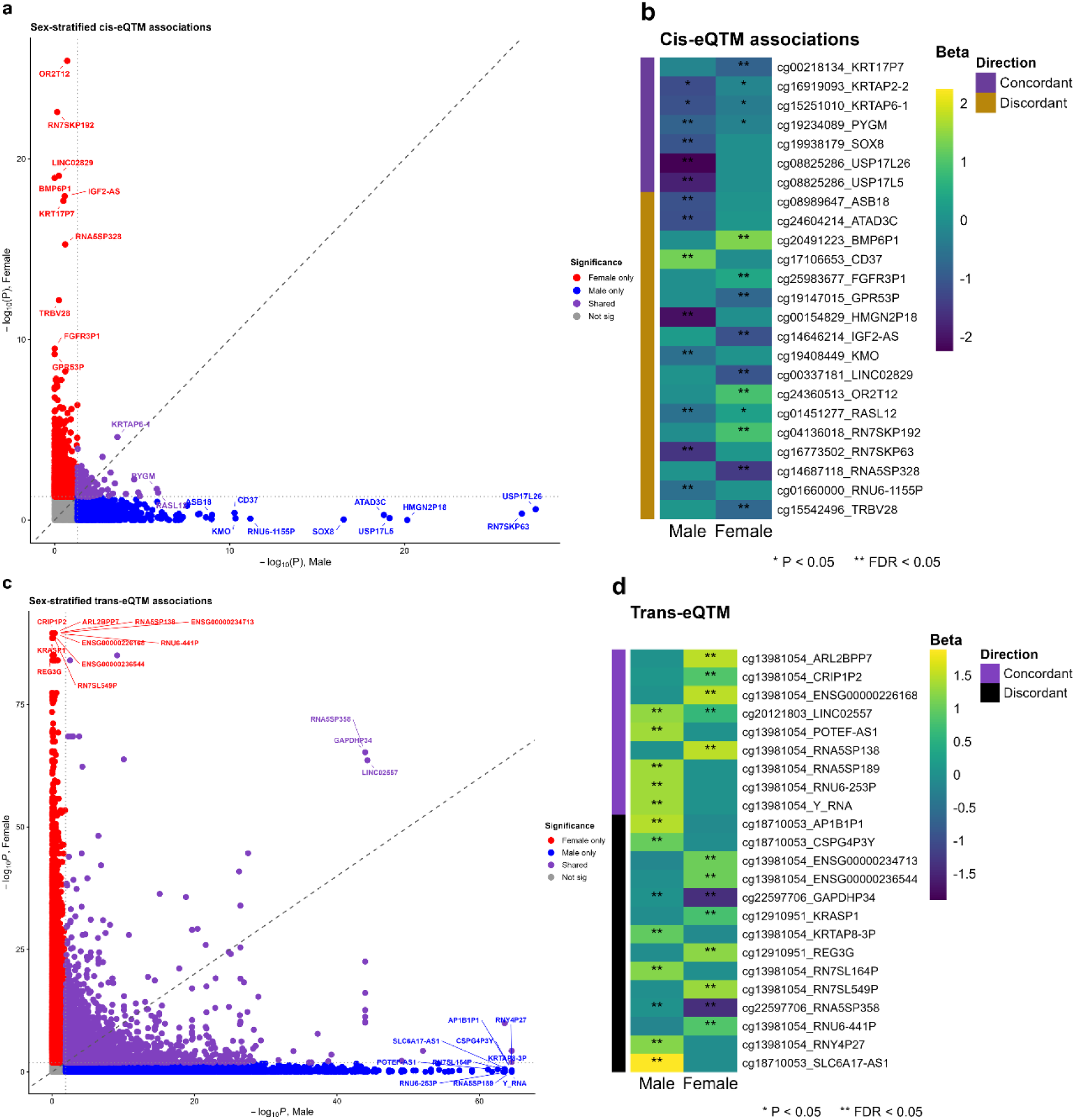
Sex-stratified cis- and trans-eQTM associations. a) Scatterplot of −log10(*P*) values for CpG– gene cis-eQTM pairs in males (x-axis; n = 115) and females (y-axis; n = 81). Dotted lines indicate *P* = 0.05, and the dashed diagonal line indicates equal statistical evidence in both sexes. Red indicate associations significant in females only; blue, males only; purple, both sexes; and gray, neither sex. Top genes in each category are labeled. b) Heatmap of male- and female-specific β coefficients for the labeled cis-eQTM pairs in A. Colors represent β coefficients centered at zero, and the sidebar indicate concordant (same direction) or discordant (opposite direction) effects between sexes. c) Scatterplot of −log10(*P*) values for CpG–gene trans-eQTM pairs in males (x-axis; n = 55) and females (y-axis; n = 81). Dotted lines indicate *P* = 0.013, and the dashed diagonal line indicates equal statistical evidence in both sexes. Red indicates associations significant in females only; blue, males only; purple, both sexes; and gray, neither sex. Top genes in each category are labeled. d) Heatmap of male- and female-specific β coefficients for the labeled trans-eQTM pairs in C. Colors represent β coefficients centered at zero, and the sidebar indicates concordant (same direction) or discordant (opposite direction) effects between sexes. *P* < 0.05; **FDR < 0.05. Models were fitted separately by sex and adjusted for age, site, plate, PC1, and PC2.

#### Male-specific model

In the male-stratified analysis, 10,055 cis-eQTMs were identified at P < 0.05, of which 59 remained significant at FDR < 0.05 (Fig. 2a, Supplementary Table S2e). In addition, 1,139,578 trans-eQTMs were significant at FDR < 0.05 (Fig. 2b), 947549 of which were identified only in the male model (Supplementary Table S2f). Ranked by P value, the top cis-eQTM genes were *USP17L26, RN7SKP63, HMGN2P18, USP17L5 and ATAD3C*, and the top trans-eQTM genes were *RNY4P27, KRTAP8-3P, AP1B1P1, CSPG4P3Y* and *SLC6A17-AS1* (Fig. 3a-d).

#### Sex-interaction model

In the sex × CpG interaction analysis, 4,678 cis-eQTMs were identified at P < 0.05, of which 72 remained significant at FDR < 0.05 (Fig. 2a, Supplementary Table S2g). In addition, 113,305 trans-eQTMs showed a significant sex × CpG interaction at FDR < 0.05 (Fig. 2b, Supplementary Table S2h). Ranked by interaction P value, the top cis-eQTM genes were *USP17L26, HMGN2P18, USP17L5, BMP6P1* and *SOX8* (Supplementary Table S5a). The most significant sex-dependent trans-eQTMs involved *SCART1, MRPL42P6, CLCN6* and *HTR1F* (interaction FDR < 0.05; Supplementary Table S5b). Methylation was associated with SCART1, *HTR1F* and *MRPL42P6* expression in males only (male FDR < 0.045), whereas *CLCN6* showed significant associations in female only participants with asthma (P < 4.2 × 10⁻׈; Supplementary Table S5b).

### Shared and Sex-Specific eQTM Associations

Comparison of cis- and trans-eQTMs across the combined, female-specific, male-specific, and sex-interaction models revealed substantial regulatory heterogeneity, with fewer than 0.02% of associations shared across all four models (Fig. 4a, b). The combined, female-specific, male-specific, and sex-interaction models identified 5,821, 3,808, 5,254, and 5,604 unique cis-eQTMs, respectively, yet only 14 cis-eQTMs were shared across all four models. Model specificity was even more pronounced for trans-eQTMs: female-specific and male-specific analyses identified 823,286 and 947,549 unique trans-eQTMs, respectively, whereas only 379 trans-eQTMs (<0.02%) were common to all models. A total of 93 cis-eQTMs and 67 trans-eQTMs were consistently identified across the female-specific, male-specific, and sex-interaction models but were absent from the combined sex-adjusted analysis, highlighting the ability of sex-aware approaches to detect regulatory relationships that would otherwise remain undetected. The top 30 cis-eQTM and trans-eQTM CpG-gene pairs are shown in Tables 2 and 3, respectively.

**Figure 4.**
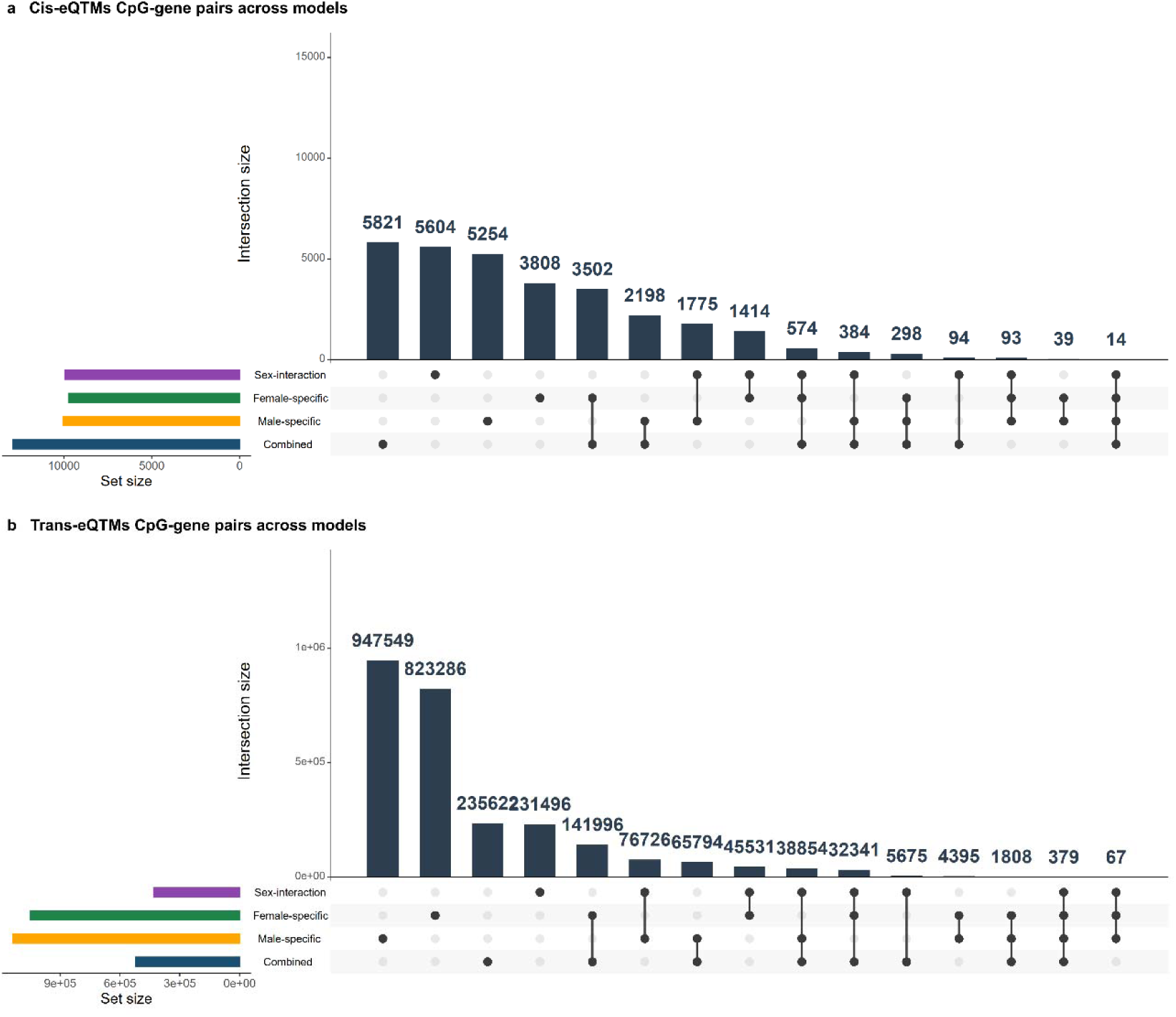
Upset plots illustrating the overlap of asthma specific cis (top) and trans (bottom) eQTMs across four analytical models: Combined, Female Specific, Male Specific, and Sex Interaction. Each bar represents the number of CpG–gene pairs uniquely or jointly identified b each model. Model specific colors correspond to Combined (blue), Female Specific (green), Male Specific (orange), and Sex Interaction (purple). Only asthma specific eQTMs (significant in asthma but not in controls) are included. Cis and trans results are displayed as separate panels to highlight differences in local versus distal epigenetic regulation.

**Table 2.**
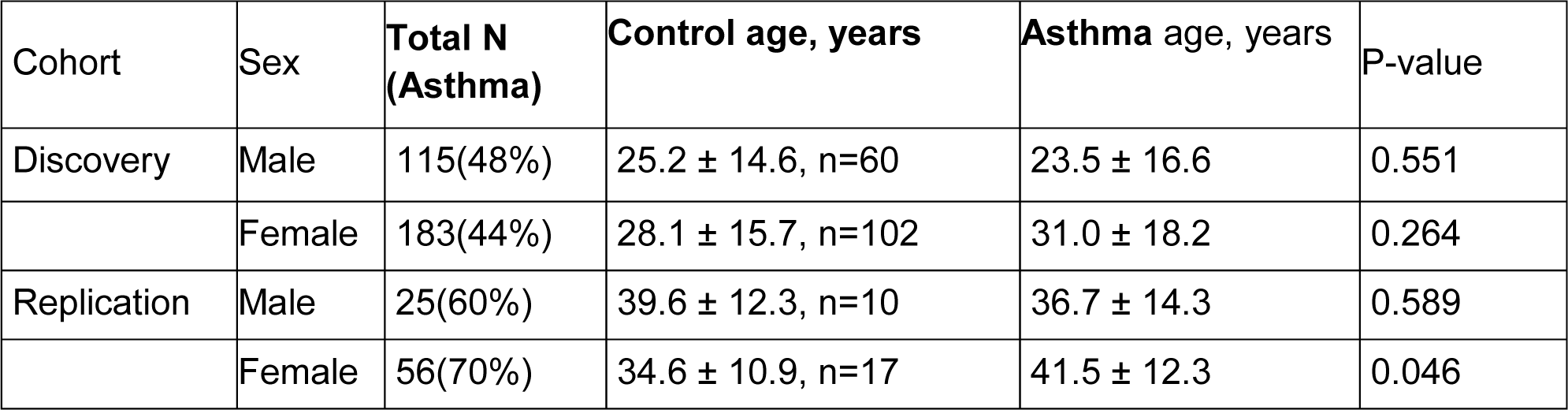
Top 30 cis-eQTM CpG–gene pairs that are significant in the sex-interaction, male-specific, and female-specific models but not in the combined model in subjects with asthma.

**Table 3.**
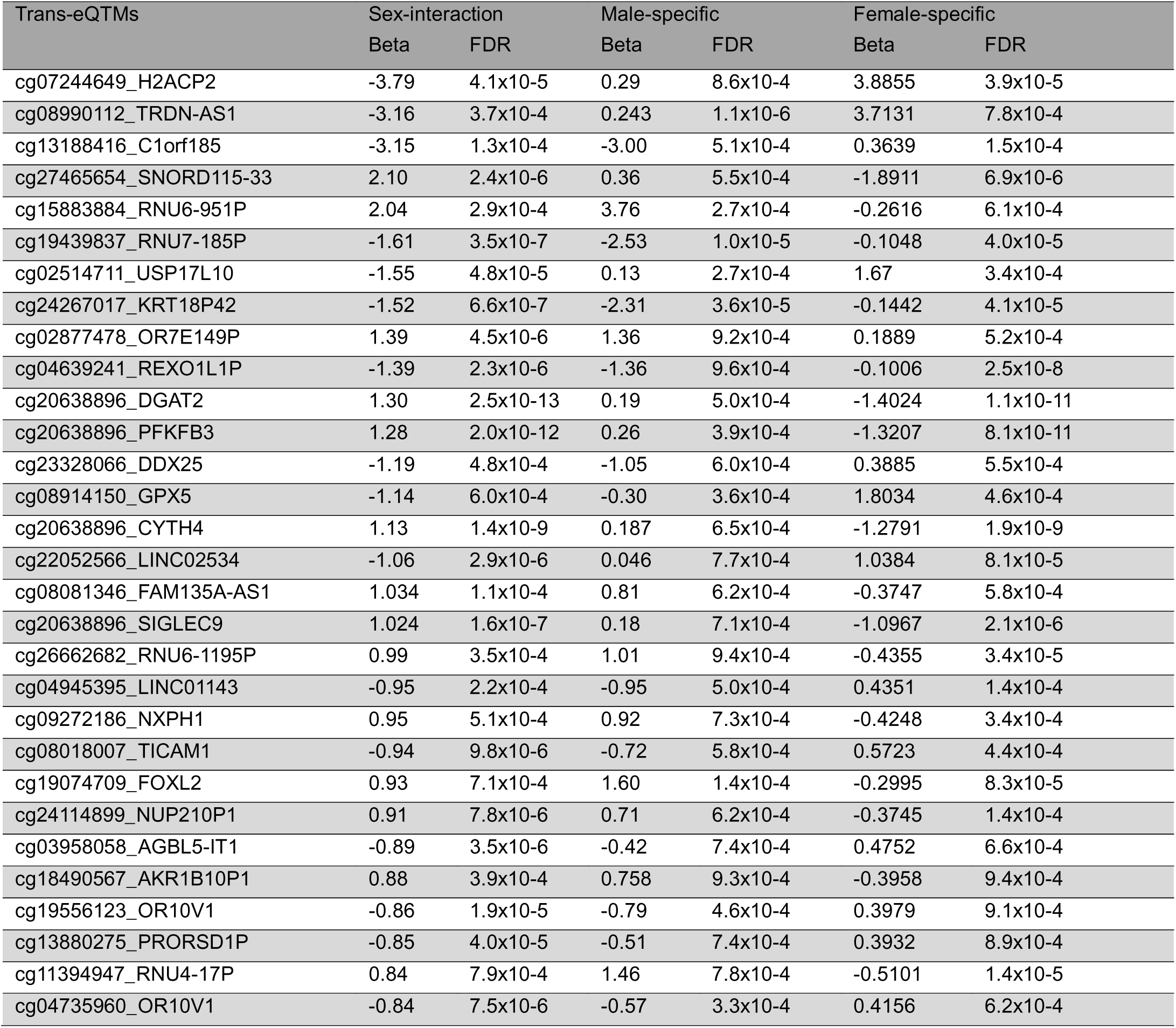
Top 30 trans-eQTM CpG–gene pairs that are significant in the sex-interaction, male-specific, and female-specific models but not in the combined model in subjects with asthma.

We next asked whether eQTM target genes were differentially expressed between participants with asthma and healthy controls. In the sex-combined model, 389 cis-eQTMs (3.0%) and 12,829 trans-eQTMs (2.5%) involved genes that were differentially expressed by asthma status (Supplementary Table S3a, b). Ranked by eQTM P-value, the strongest of these associations involved *SSTR2, MSS51, C12orf54, AKR1B1, KRT15 and GFRA1* in cis, and *CTSG, CYP2A7, S1PR1, NTRK2* and *NTRK1* in trans. Differentially expressed eQTM genes also included genes with established roles in asthma and airway inflammation, including *IL18RAP, EGR1, RGS16, MUC17* and *SPON2* among cis-eQTMs, and *IL1RL1, TSLP, NPSR1, CLCA1* and *PTGDR2* among trans-eQTMs.

We then asked whether eQTMs that were significant in the male- or female-specific analyses, and that also showed a significant sex × methylation interaction, involved genes differentially expressed by asthma status within the same sex. To do this, we intersected these sex-specific, sex-interacting cis- and trans-eQTMs with the sex-stratified differential expression results. Of the 4,678 cis and 113,305 trans sex-interacting eQTMs, 662 (14.2%) and 4,006 (3.5%), respectively, involved genes that were differentially expressed in males or females with asthma (Fig. S1a and S2b). Among cis-eQTMs, the top differentially expressed genes in males were *SCART1*, *MRPL42P6*, *HTR1F*, *IGHG4*, *BCAP29*, *COL14A1*, *THY1*, *COMP*, *IFT74* and *TRPC6*, and in females *CLCN6*, *ANGPTL7*, *MIR22HG*, *WSB1*, *POLQ*, *LDHAL6A* and *RPS11P5*. Among trans-eQTMs, the top genes in males were *MTCO3P12*, *EEF1A2*, *FCRL2*, *CNR2*, *S1PR1*, *COL4A3*, *IGHG4* and *CYP1B1-AS1*, and in females *C4BPA*, *CYP2A7*, *HS3ST4*, *SHC4* and *IL31RA*. These genes span immune signaling (*IGHG4*, *FCRL2*, *CNR2*, *C4BPA*, *IL31RA*), extracellular matrix (*COL14A1*, *COMP*, *COL4A3*, *THY1*) and ion-channel or receptor signaling (*TRPC6*, *CLCN6*, *HTR1F*). This suggests that genes with sex-dependent methylation expression associations are also differentially expressed in asthma within the corresponding sex.

### Replication of eQTMs in bronchial airway epithelium replication cohort

To assess whether eQTMs identified in nasal epithelium extend to the lower airway, we tested replication in an independent bronchial airway epithelial dataset with paired DNA methylation and RNA-seq data. Replication was evaluated for all four models (sex-combined, female, male and sex × CpG interaction) and was defined as a concordant direction of effect with nominal significance in the replication dataset, among asthma-specific discovery pairs (Methods). In total, 451 CpG–gene pairs replicated: 3 cis and 96 trans-eQTM in the sex-combined model, 1 cis and 84 trans in the female model, 9 cis and 216 trans-eQTM in the male model, and 2 cis and 41 trans-eQTM in the interaction model (Fig. 5a, Supplementary Fig.S3-6).

**Figure 5.**
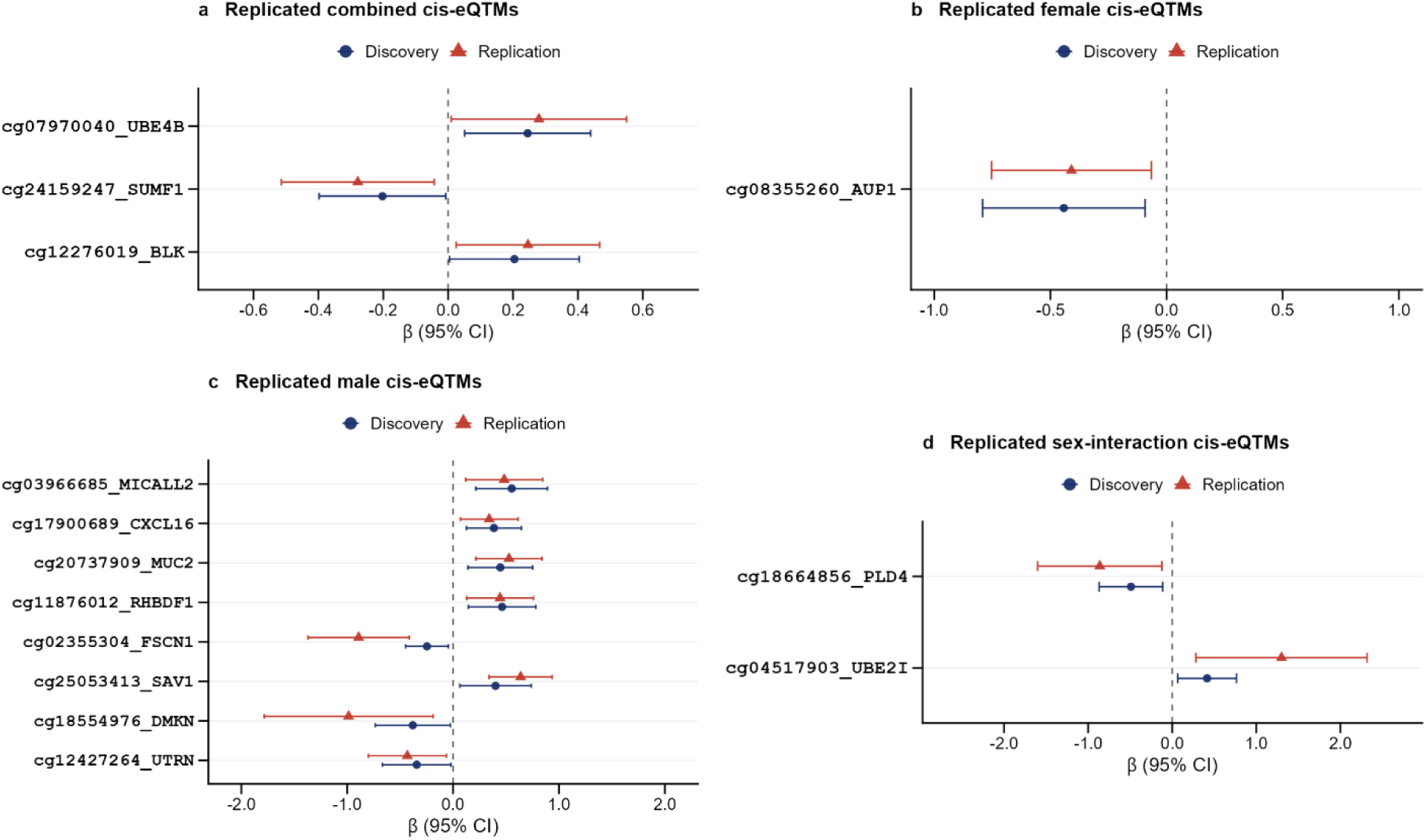
Replicated cis-eQTM associations across sex-stratified models. Forest plots show β estimates and 95% confidence intervals (horizontal bars) for replicated cis-eQTM pairs identified after exclusion of associations significant in control subjects, in a) the combined model, b) the female-specific model, c) the male-specific model, and (d) the sex-interaction model. Each row represents one CpG– gene pair, ordered by discovery p-value (most significant at top). The vertical dashed line marks β = 0. Trans-eQTM pairs were required to pass FDR < 0.05 in discovery, be absent from control-subject associations (p < 0.05), and achieve nominal replication (p < 0.05) with concordant effect direction in the independent Replication dataset. Abbreviations: β, standardized regression coefficient; CI, confidence interval; trans-eQTM, cis expression quantitative trait methylation locus.

In the sex-combined model, the replicated cis-eQTM genes were UBE4B, SUMF1 and BLK, whereas the replicated trans-eQTMs were dominated by immune and inflammatory genes, including *TNF, IRF8, CD86, CXCL11, SLAMF7, TNFSF13B, CCL26, FCGR3B, S100A9* and *NLRC3*. Most of these genes were inversely associated with methylation at a single CpG in both datasets (Fig. 5a and Fig.6a). In the female model, the cis-eQTM gene AUP1 replicated, and the replicated trans genes included the type 2 inflammatory genes *IL4, IL18R1* and *POSTN*, together with *SHH, IRF8, TNF, TNFSF13B, FCGR3B, S100A9* and *DNASE1L3*, all with concordant effect directions across tissues (Fig. 5b and Fig.6b). The male model yielded the most replicated associations, with nine cis-eQTM genes (*MICALL2, CXCL16, AUP1, MUC2, RHBDF1, FSCN1, SAV1, DMKN* and *UTRN*). Among the replicated trans-eQTMs, methylation at a single CpG near RHBDF1 was associated with the expression of *ID3, PLK1, TRIP10, FUT3* and *EPHA1* (Fig. 5c and Fig.6c). The two replicated cis-eQTM genes, *PLD4* and *UBE2I*, showed stronger effects in bronchial than in nasal epithelium. Replicated trans-eQTMs included *ATAD3C, SLC38A11, MANF, LDB2, MS4A1, PON3, VASH1, LTB4R and MICA*, and *HOMER2* expression was associated with three distinct CpGs (Fig. 5d and Fig.6d).

Among replicated trans-eQTMs, only nine genes were also differentially expressed between asthma cases and controls within a single sex. These comprised PON3, VASH1, MS4A2, HOMER2, PLD4, and CHDH in females, and AMY2B, FMO1, MICA, and CHDH in males. Notably, CHDH was the sole gene differentially expressed in both sexes.

### A subset of replicated CpGs is associated with both nearby and distant genes

Six of the CpGs with replicated cis-eQTMs were also associated with distant genes in trans within the same model, in both the discovery and replication datasets, whereas the remaining CpGs showed cis associations only (Fig. 7a and b )). The most extensive pattern was observed in males.

**Figure 6.**
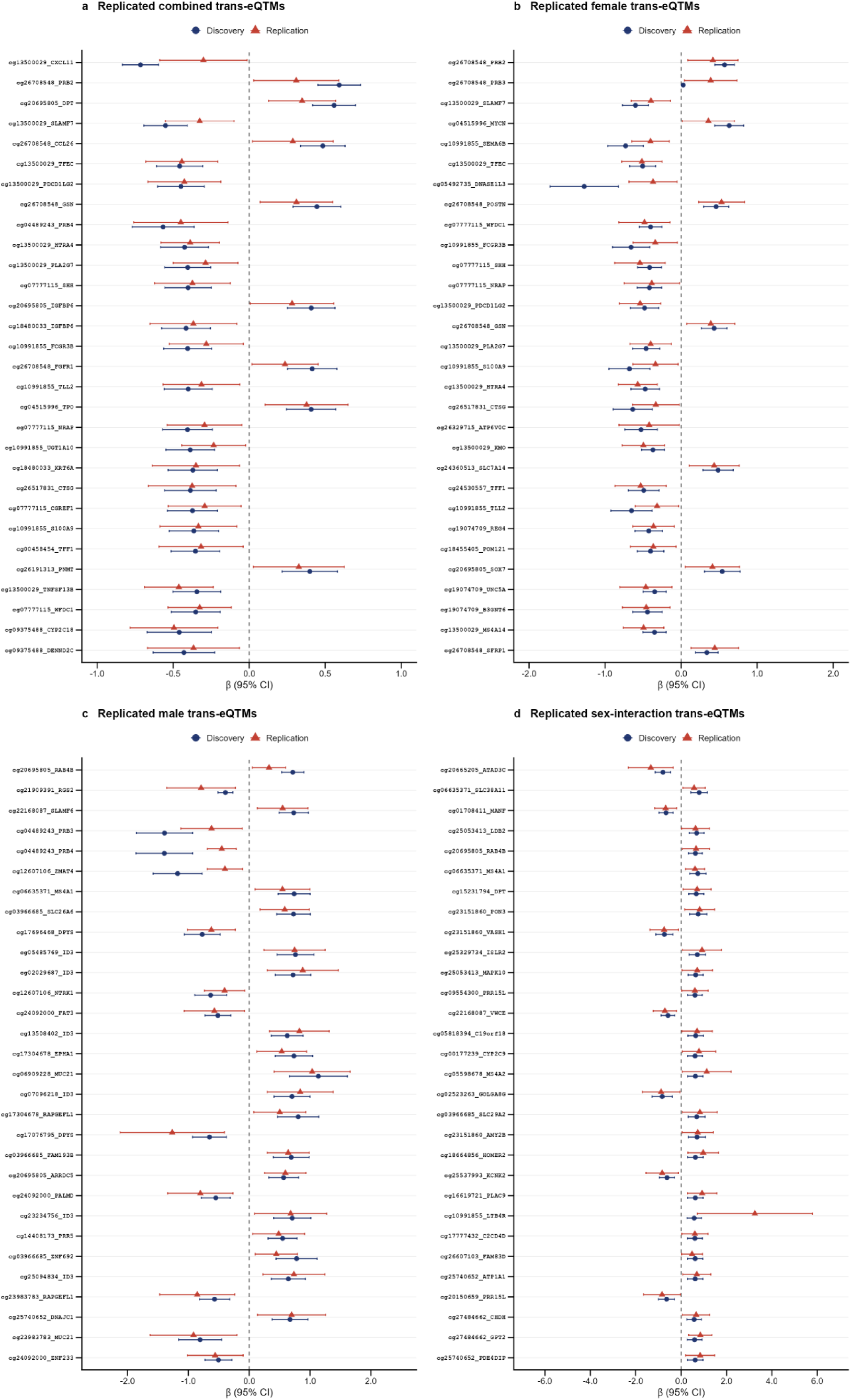
Replicated trans-eQTM associations across sex-stratified models. Forest plots show β estimates (points) and 95% confidence intervals (horizontal bars) for replicated trans-eQTM pairs identified after exclusion of associations significant in control subjects, in **a)** the combined model, **b)** the female-specific model, **c)** the male-specific model, and **d)** the sex-interaction model. Each row represents one CpG–gene pair, ordered by discovery p-value (most significant at top). The vertical dashed line marks β = 0. Trans-eQTM pairs were required to pass FDR < 0.05 in discovery, be absent from control-subject associations (FDR < 0.05), and achieve nominal replication (p < 0.05) with concordant effect direction in the independent replication dataset. The top 30 pairs by discovery p-value are shown per model. Abbreviations: β, standardized regression coefficient; CI, confidence interval; trans-eQTM, trans expression quantitative trait methylation locus; FDR, false discovery rate.

**Figure 7.**
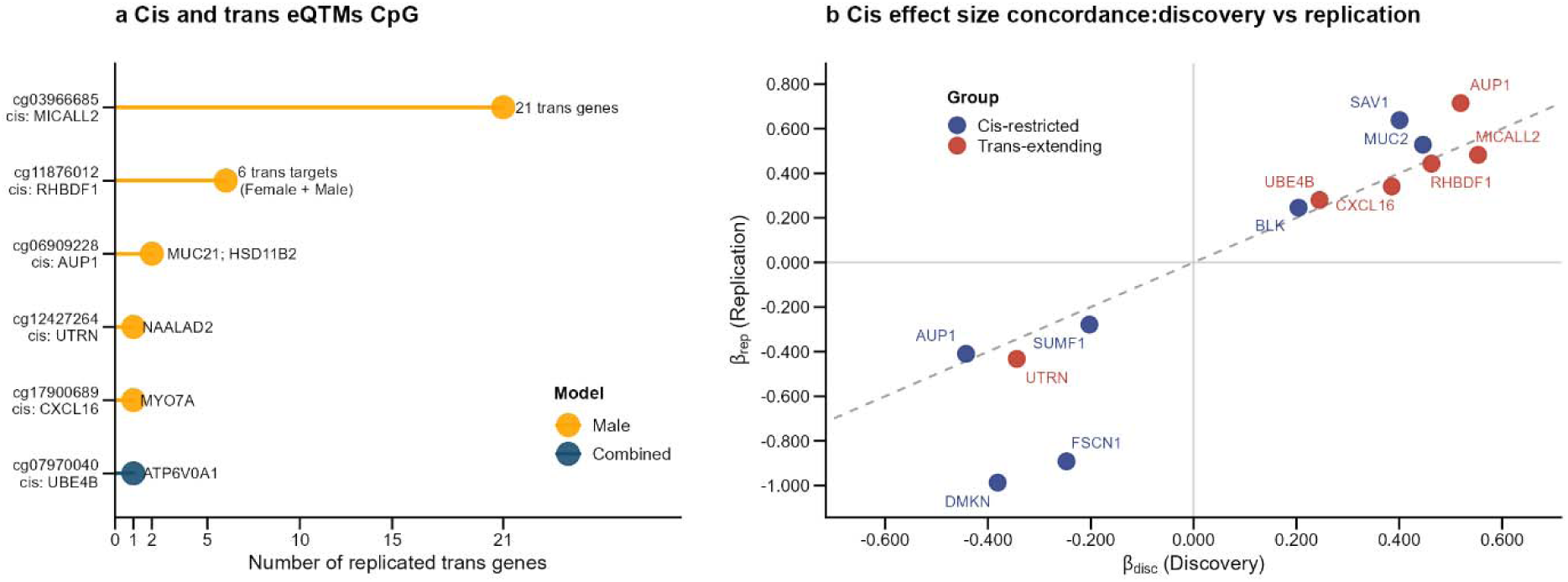
Cis and trans-eQTM CpG sites and their trans-regulated genes. a) Number of replicated trans-eQTM genes for each of the six cis-and-trans CpGs. Y axis showing Each CpG is with cis-eQTMs gene in y axis and the number of replicated trans genes. **b)** Concordance of cis effect sizes between discovery, x-axis and Replication, y-axis dataset for all 14 replicated cis eQTM pairs. Each point represents one CpG–gene pair, labelled by gene symbol. The dashed diagonal line indicates perfect concordance. Colors indicate CpG classification: red = Cis and trans-eQTMs (n = 6), navy = Cis-eQTMs only (n = 7). Concordant effect directions across datasets confirms replication.

The DNA methylation site cg03966685 associated with *MICALL2* in cis was associated in trans with multiple genes, including the chromatin regulators *HDAC7, HDAC10* and *KAT2A* and the RNA-processing genes *DGCR8, MOV10* and *PABPN1*. The DNA methylation site cg11876012 with *RHBDF1* in cis was associated in trans with *ID3, PLK1, TRIP10, FUT3* and *EPHA1* in males and, in the female model, was inversely associated with *AKR1C3* in both datasets. Two further male-specific CpGs linked a cis gene to a distant gene: *CXCL16* with *MYO7A*, and *UTRN* with *NAALAD2*. In the sex-combined model, the CpG associated with *UBE4B* in cis was also associated with *ATP6V0A1* in trans, and in the interaction model the CpG associated with *PLD4* in cis was positively associated with *HOMER2* in trans. These findings identify a small set of CpGs whose methylation is reproducibly linked to the expression of both a nearby gene and distant genes, predominantly in males. Whether these relationships reflect direct regulation will require functional studies.

Overall, a subset of the asthma-associated methylation–expression relationships identified in nasal epithelium replicated in bronchial epithelium, including associations with the immune and type 2 inflammatory genes *TNF, IRF8, IL4* and *POSTN*, suggesting that epigenetic regulation of gene expression is shared across the upper and lower airway in asthma.

### Replicated sex-specific trans-eQTMs implicate asthma-related pathways

To characterize the biological functions of replicated methylation-associated trans-regulatory networks, we performed pathway enrichment analysis of replicated trans-eQTM target genes from each model and integrated significantly enriched pathways (FDR < 0.05) into bipartite gene-pathway networks (Figs. 6 and 8). Hub genes were defined as targets participating in two or more enriched pathways. Across all models, the networks comprised 98 genes, 51 pathways, and 202 gene-pathway edges, with 47 hub genes. Only six genes were shared between models.

**Figure 8.**
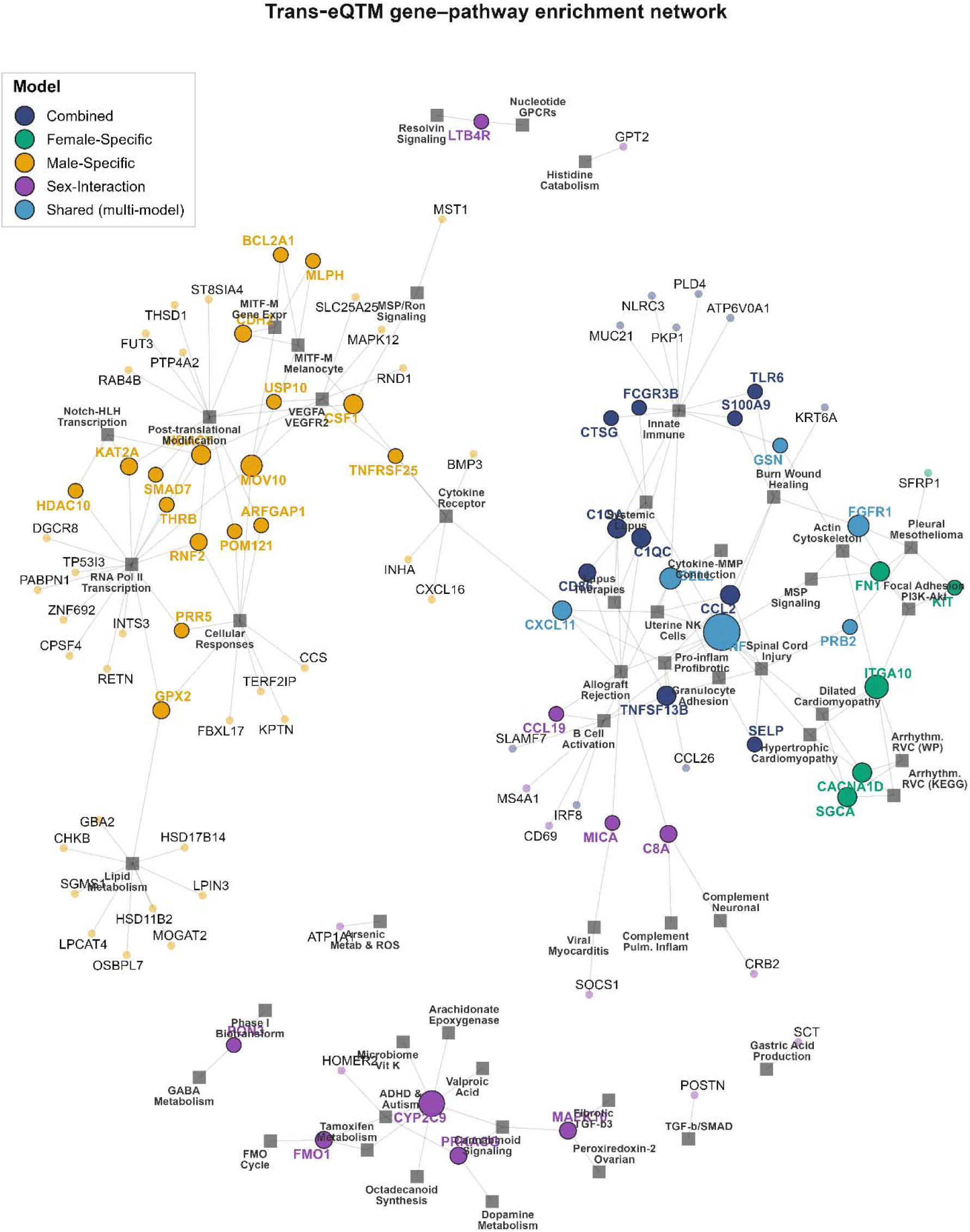
Sex-stratified trans-eQTM gene–pathway enrichment network in asthma. Bipartite network of replicated trans-eQTM associations in asthma, integrating gene–pathway memberships across four sex-stratified analytical models. Each circle represents a trans-regulated gene, colored by the model in which it was identified: navy (combined), green (female-specific), orange (male-specific), and purple (sex-interaction); genes replicated across multiple models are shown in blue (shared). Circle size is proportional to the number of enriched pathways connected to each gene (pathway degree). Hub genes (connected to ≥2 pathways) are displayed with bold labels. Each square node represents an enriched biological pathway, with size proportional to the number of connected genes. Edges connect genes to their associated pathways. The network comprises n genes, n pathways, and N gene–pathway edges in total. Abbreviations: trans-eQTM, trans-expression quantitative trait methylation locus; eQTM, expression quantitative trait methylation.

In the sex-adjusted combined model, 25 genes mapped to ten enriched pathways, primarily involving innate immunity, inflammatory signaling, and B-cell regulation. Thirteen genes were hubs, with TNF showing the highest connectivity in the combined network, followed by *C1QA, C1QC, CCL2,* and *TNFSF13B*. Other hubs included *CD86, CTSG, FCGR3B, S100A9, SELP*, and *TLR6.* These findings indicate that replicated trans-eQTM targets in the combined model converge predominantly on immune and inflammatory pathways.

In the female-specific model, 11 genes mapped to ten enriched pathways, of which eight were hubs. *ITGA10* was the most connected female-specific gene, participating in six pathways, followed by *CACNA1D* and *FN1*. Enriched pathways predominantly involved actin cytoskeleton regulation, focal adhesion, and PI3K-Akt signaling. *ITGA10, FN1*, and *FGFR1* connected multiple structural and signaling pathways, whereas shared genes such as *TNF* and *SELL* connected immune-related pathways. Notably, *TNF*, which was shared between the combined and female-specific models, was the most highly connected gene across the overall network, participating in 12 pathways. Together, these results highlight female-specific trans-regulatory networks involving cytoskeletal organization, cell adhesion, and associated signaling processes.

The male-specific network was the largest, comprising 50 genes mapped to ten enriched pathways, with 17 hub genes. Enriched pathways predominantly involved RNA polymerase II transcription, post-translational protein modification, cellular stress-responses to stimuli, and growth-factor and cytokine signaling. *MOV10* was the most connected male-specific gene, participating in five pathways, followed by *CSF1* and *HDAC7*, each participating in four pathways. Additional hubs included *CDH2, GPX2, KAT2A*, and *RNF2. CXCL11* was shared between the combined and male-specific models and connected cytokine- and immune-related pathways. Overall, the male-specific network was characterized primarily by transcriptional regulation and cellular-response pathways.

The sex-interaction model identified 18 genes mapping to 24 enriched pathways, the largest number of pathways among the four models, with nine hub genes. *CYP2C9* was the most connected gene, participating in seven pathways, including arachidonate epoxygenase and linoleic acid-derived lipid mediator pathways. Other hubs included *C8A, FMO1, MAPK10*, and *PRKACG*, each participating in three pathways, while *CCL19, LTB4R, MICA,* and *PON3* participated in two pathways. Collectively, these genes mapped to lipid mediator metabolism, complement-associated inflammation, GPCR signaling, and metabolic pathways, revealing regulatory processes that identified specifically from modeling CpG × sex interactions.

Collectively, replicated trans-eQTM targets revealed a shared inflammatory core alongside distinct sex-associated regulatory program. The combined model mainly identified immune-inflammatory pathways, whereas female-specific associations highlighted cytoskeletal and cell-adhesion signaling, male-specific associations emphasized transcriptional and cellular-response processes, and sex-interaction associations identified lipid mediator, metabolic, and complement-related pathways. These findings demonstrate that sex-stratified and sex-interaction trans-eQTM analyses reveal regulatory networks that are not fully captured by sex-adjusted models.

## Discussion

In our study, integrating asthma-associated DNA methylation with airway epithelial gene expression revealed distinct methylation expression architectures in females and males. Common between the combined and sex-stratified analyses was limited for differentially methylated CpGs and even more limited for eQTMs. A substantial part of this sex-dependent regulatory eQTM associations was therefore did not identified when sex was used as a covariate^30,31^. Sex-dependent associations were observed for both cis- and trans-eQTMs. A subset replicated in bronchial epithelium, and a smaller proportion involved genes that were differentially expressed in asthma. Together, these findings suggest that sex influences not only which CpG sites are associated with asthma, but also how DNA methylation variation at these loci relates to airway gene expression.

Asthma-associated DNA methylation patterns differed substantially by sex. Overlap between sex-combined and sex-stratified analyses was limited, with female-associated CpGs mainly showing hypomethylation, whereas male-associated CpGs were more often hypermethylated and showed stronger effects. Previous studies have reported widespread autosomal sex differences in DNA methylation in adults^32^ and children^33^,as well as sex-specific methylation changes associated with asthma^3^. Our findings extend these observations to the airway epithelium, implicating sex-dependent epigenetic regulation as a contributor to asthma-associated molecular heterogeneity. Sex-stratified analyses identified numerous trans-eQTMs that were not detected in the combined model, indicating that sex-dependent methylation expression relationships may remain unidentified when sex is used as a covariate^31,34^. Our replicated methylation-by-sex interaction eQTMs provide direct evidence of sex-dependent regulation to DNA methylation–gene expression association in the airway.

In our study, One CpG provides a clear example of how modeling sex solely as a covariate can obscure sex-specific methylation expression associations. DNA methylated site cg11876012 was not identified in the combined model. In males it showed a replicated cis association with *RHBDF1* and concordant trans-associations with *ID3*, *PLK1*, *TRIP10*, *FUT3* and *EPHA1*, whereas in females it was inversely associated with *AKR1C3* in both datasets. Two of the male-specific targets have been linked to airway biology. *PLK1* is required for airway smooth muscle cell proliferation^35^. *FUT3* encodes a fructosyltransferases, and previous studies have reported that epithelial fucosylation increases in allergic airway inflammation^36^ and may contribute to inflammatory cell recruitment^37^. These findings suggest that the same CpG may involve in distinct sex-specific regulatory networks in the airway.

Trans associations accounted for the majority of detected eQTMs in our analyses, consistent with a previous nasal epithelial study that identified extensive cis- and trans-eQTMs in atopic asthma and reported enrichment of replicated trans-eQTM genes in immune pathways and among asthma-associated differentially expressed genes¹ . Our findings extend these observations by demonstrating that trans-eQTMs can be sex-dependent and reproducible across airway compartments. Despite trans-eQTMs do not necessarily indicate direct regulation of distal genes, six CpGs were consistently associated with both local and distant targets across datasets. Importantly, a CpG linked to MICALL2 was associated in males with the chromatin regulators HDAC7, HDAC10, and KAT2A, resembling trans-eQTL architectures in which a single locus influences multiple distal genes¹¹.

Replicated trans-eQTMs in the combined model included several immune and inflammatory genes, such as TNF, IRF8, CD86, CXCL11, TNFSF13B, and S100A9, consistent with previous reports that airway trans-eQTMs are enriched for immune pathways^16^. These findings align with the marked immune heterogeneity observed in asthma, including type 2, pro-inflammatory non-type 2^38^ and neutrophil-associated^39^ programs. In addition, female-specific replicated targets included IL4, IL18R1 and POSTN. IL-4 is a central type 2 cytokine that drives IgE class switching^40^. Periostin, encoded by POSTN, is a biomarker of eosinophilic, type 2-associated airway inflammation^41^. The IL-18 pathway has been implicated in severe asthma^42^, and increased circulating IL-18R1 has been reported in asthma^43^. The enrichment of asthma-relevant genes among female-specific replicated eQTMs further supports the biological relevance of these sex-specific regulatory associations.

Integration with differential expression provided an additional level of prioritization by identifying eQTM targets that were also dysregulated in asthma. In the sex-combined analysis, these included several established asthma-related genes, including IL1RL1, TSLP, NPSR1, CLCA1, and PTGDR2. Sex-interaction eQTMs were more frequently linked to genes showing sex-specific differential expression, suggesting concordance between sex-dependent methylation– expression relationships and transcriptional variation. Among replicated trans-eQTM targets, PON3, VASH1, MS4A2, HOMER2, PLD4, and CHDH were prioritized in females, whereas AMY2B, FMO1, MICA, and CHDH were prioritized in males. Notably, MS4A2, which encodes the β-chain of the high-affinity IgE receptor FcεRI, has a well-established role in allergic immunity^44^. The remaining genes were prioritized as candidate asthma-related genes based on eQTM association from replicated eQTM associations across nasal and bronchial airway epithelium and asthma-associated transcriptional dysregulation.

Pathway analysis of replicated trans-eQTM targets revealed pronounced sex-specific biological heterogeneity, with only six of 98 network genes shared across analytical models. Replicated targets from the sex-combined model were enriched for innate immune and inflammatory pathways, whereas female-specific targets were enriched in cell adhesion, cytoskeletal, and PI3K-Akt signaling pathways, and male-specific targets in transcriptional, post-translational, and stress-response processes. Sex-interaction targets were associated with lipid mediator metabolism and complement-related inflammation. These findings are consistent with previous evidence that asthma is characterized by substantial molecular heterogeneity. Together with well-established sex differences in asthma prevalence and severity^2^, and the involvement of oxidative and cellular stress pathways in asthma pathogenesis^45^.Our results suggest that sex may represent an additional axis of variation in methylation-associated regulatory programs.

In addition to the observed sex-specific regulatory patterns, a subset of eQTMs replicated between nasal and bronchial epithelium. This finding is consistent with previous reports of substantial correspondence between nasal and bronchial transcriptomic profiles^19,20^, and with studies identifying reproducible asthma- and atopy-associated methylation signatures in nasal epithelium^10,46–48^. Consistent with previous studies, a subset of nasal eQTMs replicated in bronchial epithelium, supporting the existence of shared methylation–expression relationships across the upper and lower airways. However, the incomplete overlap observed underscores the distinct biology of nasal and bronchial tissues. Together, these findings support the use of nasal epithelium as an accessible surrogate for selected lower-airway regulatory processes while emphasizing the importance of tissue-specific regulation.

This study has several strengths, including the analysis of nasal epithelial cohort with replication in an independent bronchial epithelial dataset, enabling evaluation of methylation expression relationships across airway tissues. By integrating sex-combined, sex-stratified, and sex-interaction models, we distinguished shared and sex-dependent regulatory associations. The combined evaluation of differential methylation, gene expression, cis- and trans-eQTMs, differential expression, and pathway enrichment provided convergent evidence supporting the biological relevance of the identified associations.

Several limitations should also be considered. First, modest sample sizes, particularly after sex stratification, may have limited power to detect weaker sex-dependent effects. Second, we analyzed autosomal CpGs only and lacked sex-hormone measurements, so we could not assess sex-chromosome or hormonal contributions. Third, nasal and bronchial epithelium differ in cell composition, local immune exposure and baseline expression^49,50^. Some nasal associations that did not replicate may reflect genuine tissue-specific regulation rather than limited reproducibility. Replication was further constrained by differences in methylation platforms (EPIC versus 450K), which limited analyses to CpGs and genes shared across arrays and precluded evaluation of many EPIC-specific DMCs and eQTM associations. Nonetheless, a subset of associations, cis and trans-eQTMs, replicated despite these technical differences, supporting the robustness of the identified regulatory relationships. Incomplete clinical phenotyping across the retrospective GEO datasets further limited assessment of clinical heterogeneity and potential confounding factors.

Finally, the cross-sectional nature of the study limits inference regarding the temporal relationships between DNA methylation, gene expression, and asthma. Future studies integrating methylation and expression QTL data, together with longitudinal multi-ethnic cohorts and multi-site airway sampling, will be important for refining causal inference and assessing the generalizability of these findings. Single-cell multi-omic approaches and functional studies will further be needed to resolve cell-type-specific regulatory mechanisms and evaluate the biological consequences of the identified sex-dependent associations.

In conclusion, sex-stratified and sex-interaction analyses revealed a substantial sex-dependent component of airway epigenetic regulation in asthma that was not detected in conventional sex-adjusted analyses. Females and males showed distinct methylation patterns, largely non-overlapping cis- and trans-eQTMs and different downstream biological programs. A subset of these associations was reproducible across the upper and lower airways. Together, these findings underscore the importance of considering sex as a biological variable in airway epigenomic studies of asthma and suggest that sex-stratified analyses may help resolve molecular heterogeneity while informing future mechanistic and translational investigations.

## Supporting information

Suplemetary figure_Sex-specific multi-omics_in_Asthma

## Data Availability

All transcriptomics datasets used in our study are freely accessible at NCBI GEO (https://www.ncbi.nlm.nih.gov/geo/) with accession numbers: GSE250513, GSE240567, GSE85568 and GSE85567.

## Authors’ contributions

Conceptualization: TBM, EYD; Data Curation: EYD; Formal Analysis: EYD; Funding Acquisition: TBM; Investigation: EYD, TBM; Methodology: EYD, TBM; Project Administration: TBM; Resources: TBM, EYD, YG; Software: EYD; Supervision: TBM; Validation: EYD.; Visualization: EYD; Writing – Original Draft Preparation: EYD; Writing – Review & Editing: EYD, YG, BHR, AM, MK, TBM.

## Disclaimer

The funders of the study had no role in study design, data collection, data analysis, data interpretation, writing of the report, or decision to submit the report for publication.

## Consent for publication

All authors read the manuscript, agreed to its content, and declared no competing interest in relation to their work.

## Use of Large Language Models

Large language model-based tools were used for language editing and improvement of manuscript clarity during the preparation of this manuscript. All scientific content, analyses, interpretations, and conclusions were generated, reviewed, and verified by the authors.

