## Supplementary material for "Sex-stratified and interaction multi-omics analysis of DNA methylation and gene expression in asthma": Suplemetary figure_Sex-specific multi-omics_in_Asthma

[1]Precision Pulmonary Medicine Research Program, Division of Pulmonary, Critical Care, and Sleep Medicine, Department of Medicine, Indiana University School of Medicine, Indianapolis, IN, USA; [2]Indiana School of Medicine, Division of Pulmonary, Allergy, Critical Care, and Sleep Medicine, Department of Medicine, Indiana University School of Medicine, Indianapolis, IN, USA; [3]Division of Epidemiology, Department of Environmental and Public Health Sciences, University of Cincinnati College of Medicine, Cincinnati, OH, USA; [4]Department of Computer Science, University of Nevada, Las Vegas, NV, USA.

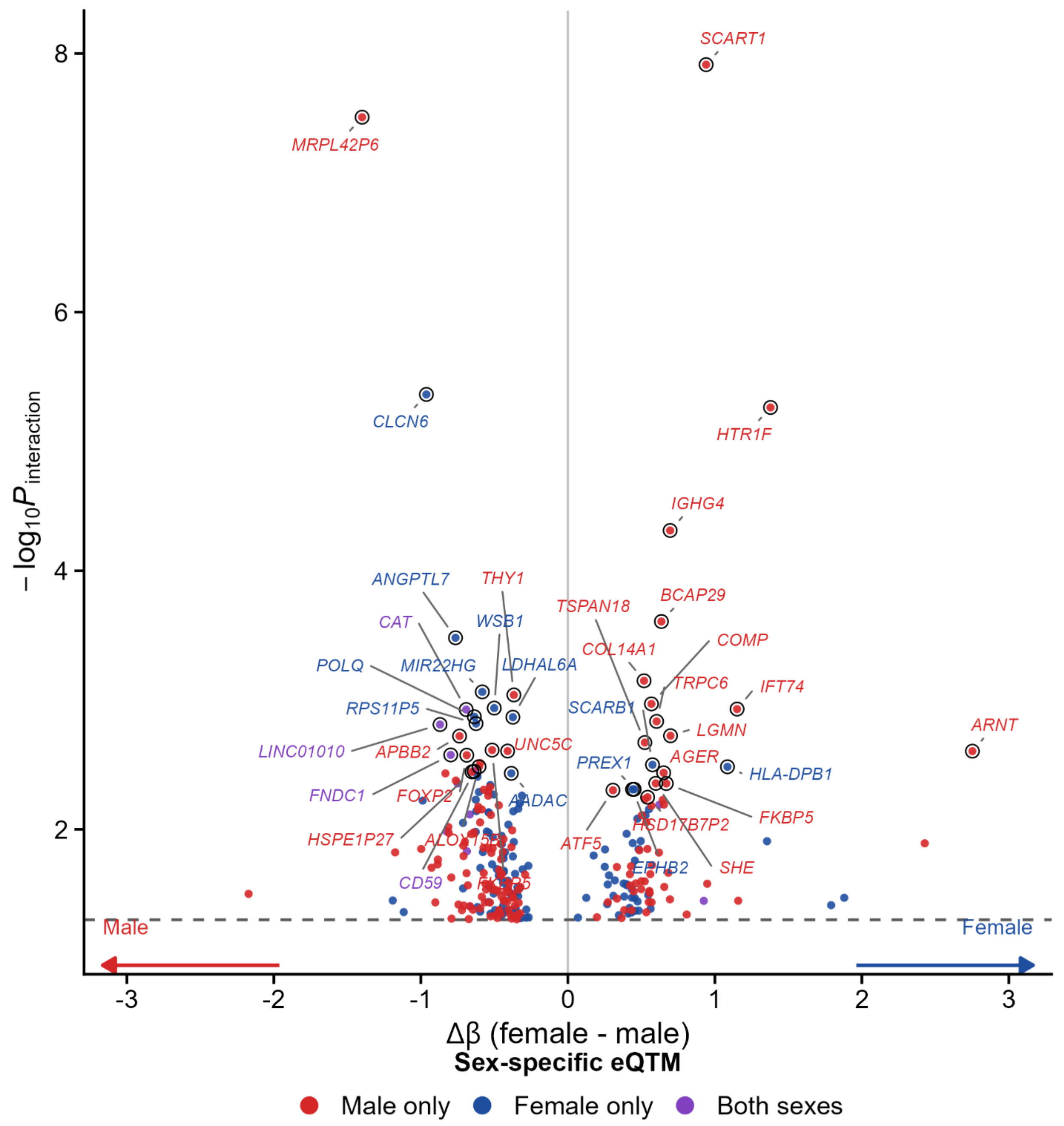


**Figure S1a**. **Sex-dependent cis-eQTMs CpG–gene pairs in participants with asthma.**

Volcano plot of the sex × CpG methylation interaction for asthma-specific cis-eQTMs whose target genes were differentially expressed in males or females with asthma (nasal epithelium). Each point is one CpG–gene pair. The x-axis shows the difference in eQTM effect size between sexes (Δβ = β_female − β_male). The y-axis shows −log10 P for the sex × methylation interaction term from a model of gene expression on CpG methylation, sex and their interaction, adjusted for covariates. Points to the right of zero have a larger (more positive) methylation–expression effect in females, and points to the left have a larger effect in males, as indicated by the arrows. Colors show the sex-stratified eQTM result: red, significant in males only; blue, significant in females only; purple, significant in both sexes (nominal $P < 0.05$ in the sex-stratified models). The dashed horizontal line marks interaction $P = 0.05$. Circled points with italic labels are the top interaction pairs on each side of zero, labelled by target gene symbol. Full statistics for all pairs are provided in Supplementary Table S3a.

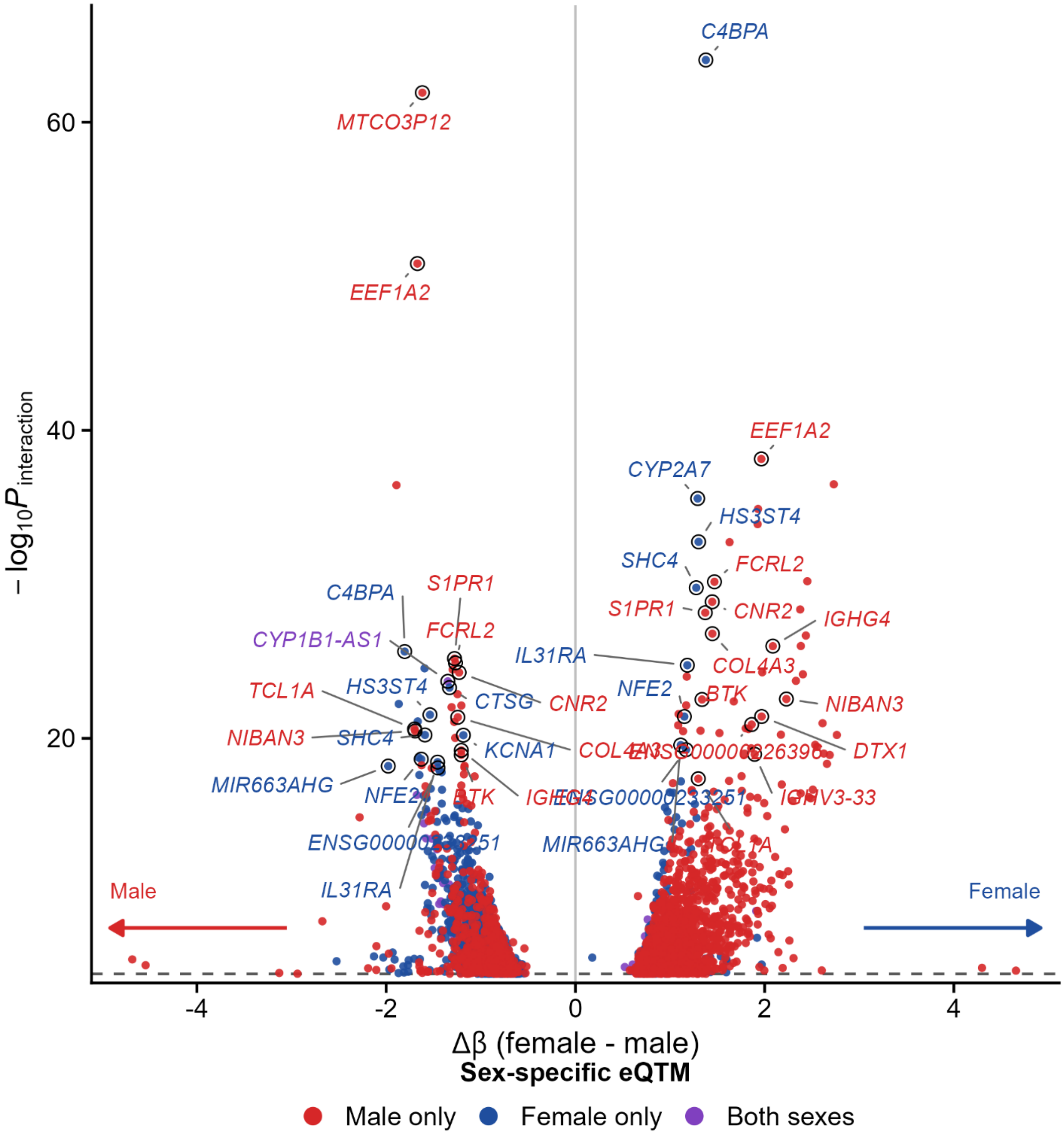


**Figure S2b. Sex-dependent trans-eQTMs CpG–gene pairs in participants with asthma.**

Volcano plot of the sex × CpG methylation interaction for asthma-specific trans-eQTMs whose target genes were differentially expressed in males or females with asthma (nasal epithelium). Each point is one CpG–gene pair. The x-axis shows the difference in eQTM effect size between sexes (Δβ = β_female − β_male). The y-axis shows −log10 P for the sex × methylation interaction term from a model of gene expression on CpG methylation, sex and their interaction, adjusted for covariates. Points to the right of zero have a larger (more positive) methylation–expression effect in females, and points to the left have a larger effect in males, as indicated by the arrows. Colors show the sex-stratified eQTM result: red, significant in males only; blue, significant in females only; purple, significant in both sexes (nominal $P < 0.05$ in the sex-stratified models). The dashed horizontal line marks interaction $P = 0.05$. Circled points with italic labels are the top interaction pairs on each side of zero, labelled by target gene symbol. Full statistics for all pairs are provided in Supplementary Table S4a.

### Replicated combined trans-eQTMs — Supplementary

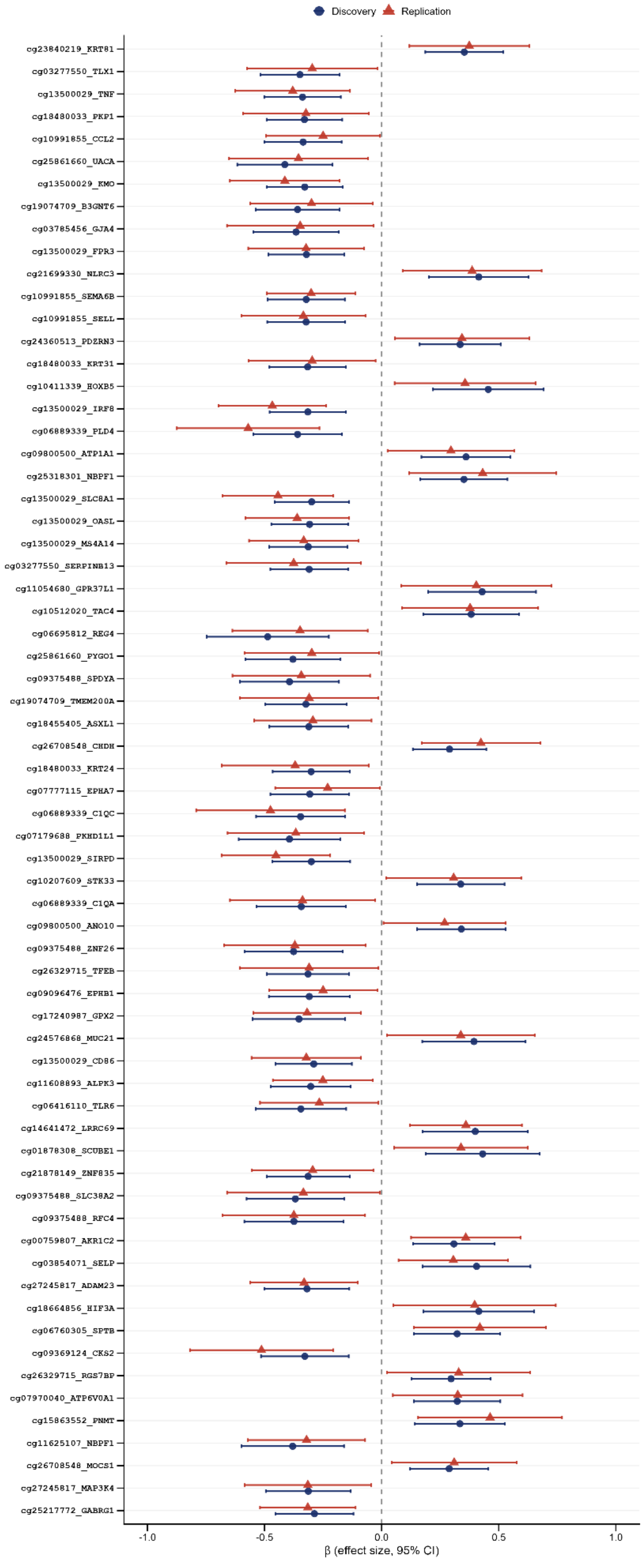

**Figure S3**. **Replicated trans-eQTM associations in the combined model.**

Forest plots show β estimates (points) and 95% confidence intervals (horizontal bars) for trans-eQTM associations identified in the combined discovery analysis and replicated in the independent replication dataset. Trans-eQTM pairs were required to meet FDR $< 0.05$ in the discovery analysis, show no significant association in control subjects (FDR $\geq 0.05$), and achieve nominal significance ($P < 0.05$) with a concordant effect direction in the replication dataset. The vertical dashed line indicates $\beta = 0$. Abbreviations: β, standardized regression coefficient; CI, confidence interval; trans-eQTM, trans expression quantitative trait methylation; FDR, false discovery rate.

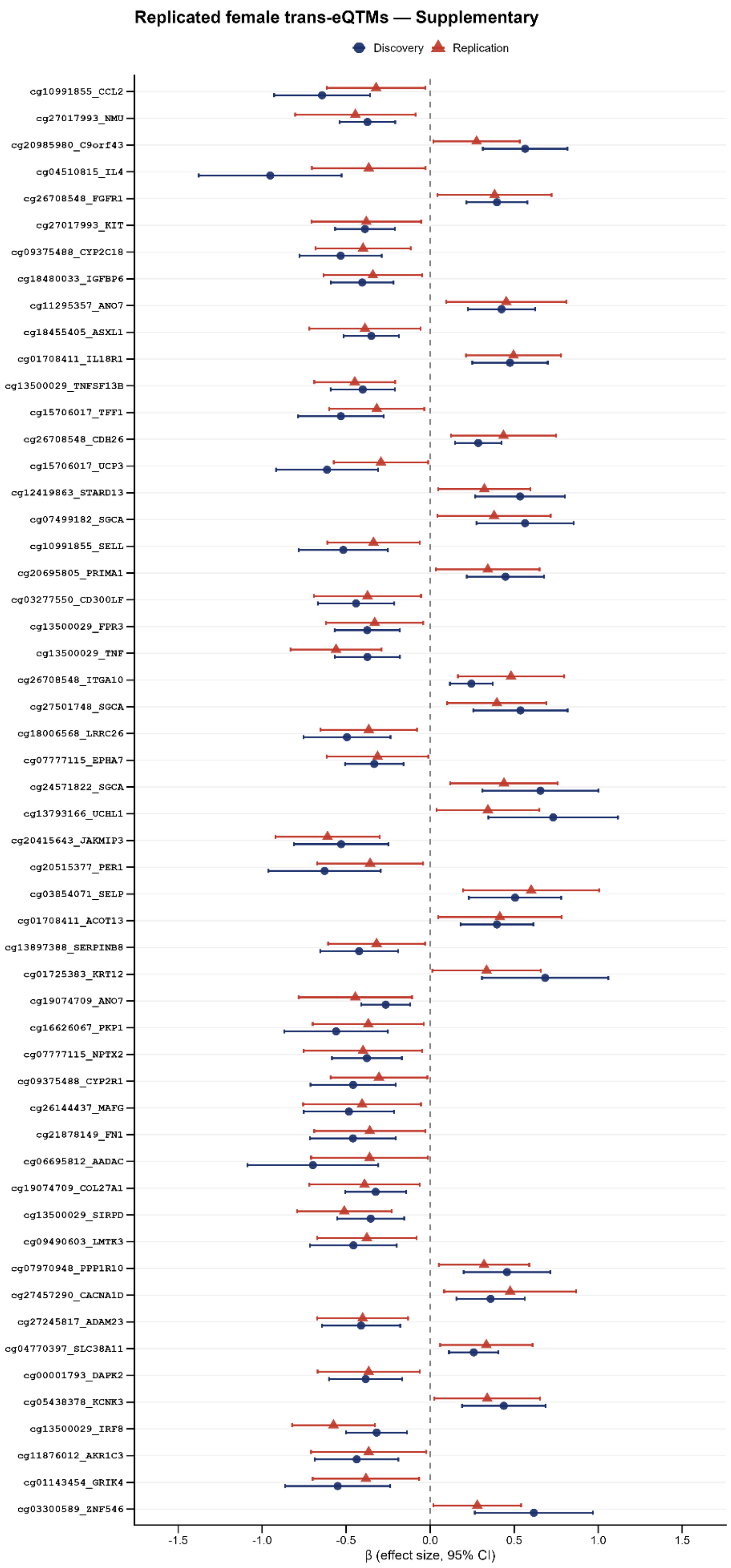
Replicated female trans-eQTMs — Supplementary
Discovery
Replication
cg10991855_CCL2
cg27017993_NMU
cg20985980_C9orf43
cg04510815_IL4
cg26708548_FGFR1
cg27017993_KIT
cg09375488_CYP2C18
cg18480033_IGFBP6
cg11295357_ANO7
cg18455405_ASXL1
cg01708411_IL18R1
cg13500029_TNFSF13B
cg15706017_TFF1
cg26708548_CDH26
cg15706017_UCP3
cg12419863_STARD13
cg07499182_SGCA
cg10991855_SELL
cg20695805_PRIMA1
cg03277550_CD300LF
cg13500029_FPR3
cg13500029_TNF
cg26708548_ITGA10
cg27501748_SGCA
cg18006568_LRRC26
cg07777115_EPHA7
cg24571822_SGCA
cg13793166_UCHL1
cg20415643_JAKMIP3
cg20515377_PER1
cg03854071_SELP
cg01708411_ACOT13
cg13897388_SERPINB8
cg01725383_KRT12
cg19074709_ANO7
cg16626067_PKP1
cg07777115_NPTX2
cg09375488_CYP2R1
cg26144437_MAFG
cg21878149_FN1
cg06695812_AADAC
cg19074709_COL27A1
cg13500029_SIRPD
cg09490603_LMTK3
cg07970948_PPP1R10
cg27457290_CACNA1D
cg27245817_ADAM23
cg04770397_SLC38A11
cg00001793_DAPK2
cg05438378_KCNK3
cg13500029_IRF8
cg11876012_AKR1C3
cg01143454_GRIK4
cg03300589_ZNF546
-1.5
-1.0
-0.5
0.0
0.5
1.0
1.5
β (effect size, 95% CI)

**Figure S4. Replicated trans-eQTM associations in the female-specific model.**

Forest plots show β estimates (points) and 95% confidence intervals (horizontal bars) for trans-eQTM associations identified in the female-specific discovery analysis and replicated in the independent replication dataset. Trans-eQTM pairs were required to meet FDR < 0.05 in the discovery analysis, show no significant association in female control subjects (FDR ≥ 0.05), and achieve nominal significance ($P < 0.05$) with a concordant effect direction in the replication dataset. The vertical dashed line indicates $\beta = 0$. Abbreviations: β, standardized regression coefficient; CI, confidence interval; trans-eQTM, trans expression quantitative trait methylation; FDR, false discovery rate.

**Replicated female trans-eQTMs — Supplementary**

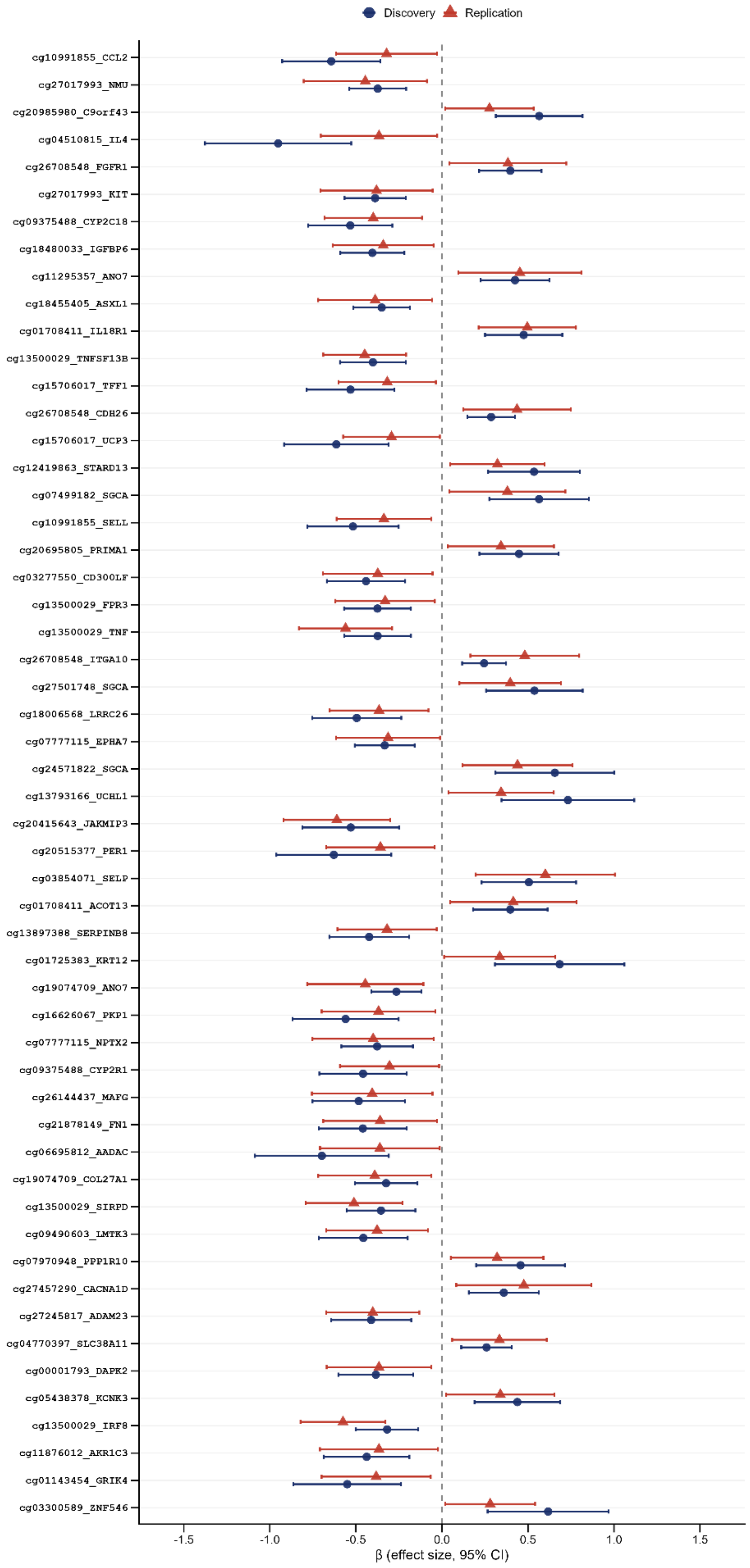

**Figure S5. Replicated trans-eQTM associations in the male-specific model.**

Forest plots show β estimates (points) and 95% confidence intervals (horizontal bars) for trans-eQTM associations identified in the male-specific discovery analysis and replicated in the independent replication dataset. Trans-eQTM pairs were required to meet FDR < 0.05 in the discovery analysis, show no significant association in male control subjects (FDR ≥ 0.05), and achieve nominal significance (P < 0.05) with a concordant effect direction in the replication dataset. The vertical dashed line indicates β = 0. Abbreviations: β, standardized regression coefficient; CI, confidence interval; trans-eQTM, trans expression quantitative trait methylation; FDR, false discovery rate.

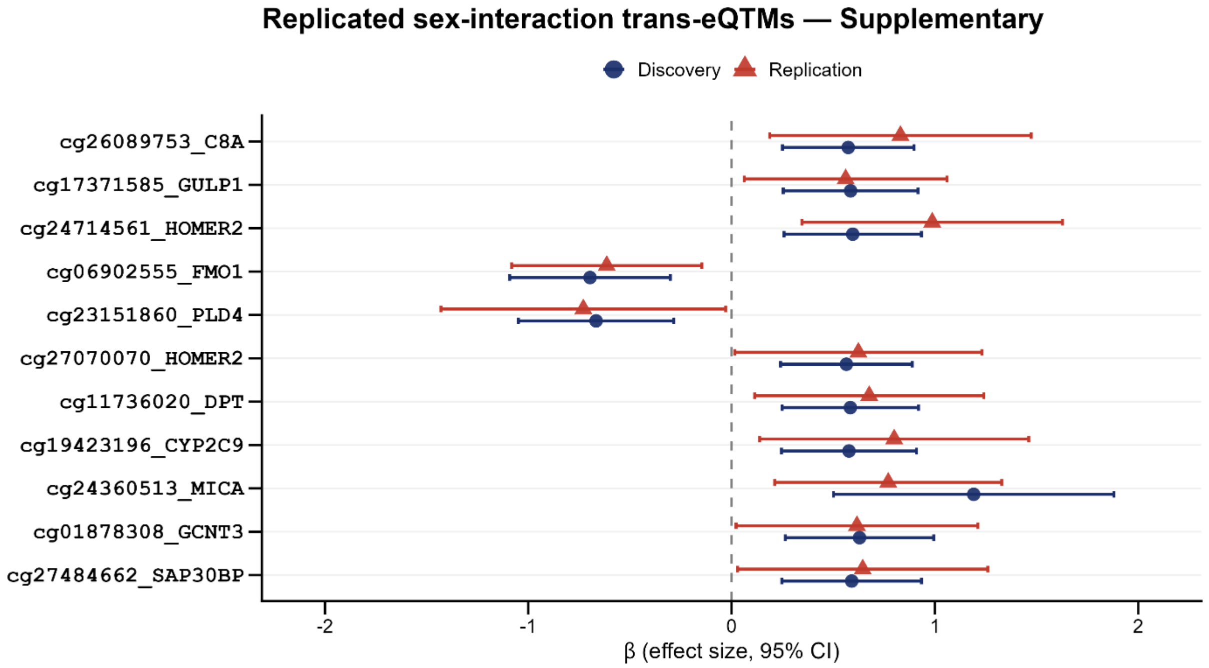


**Figure S6. Replicated trans-eQTM associations in the sex-interaction model.**

Forest plots show β estimates (points) and 95% confidence intervals (horizontal bars) for trans-eQTM associations identified in the sex-interaction discovery analysis and replicated in the independent replication dataset. Trans-eQTM pairs were required to meet FDR $< 0.05$ in the discovery analysis, show no significant association in control subjects (FDR $\geq 0.05$), and achieve nominal significance ($P < 0.05$) with a concordant effect direction in the replication dataset. The vertical dashed line indicates $\beta = 0$. Abbreviations: β, standardized regression coefficient; CI, confidence interval; trans-eQTM, trans expression quantitative trait methylation; FDR, false discovery rate.
